# Care Models for Long COVID : A Rapid Systematic Review

**DOI:** 10.1101/2021.11.17.21266404

**Authors:** Simon Décary, Michèle Dugas, Théo Stefan, Léa Langlois, Becky Skidmore, Anne Bhéreur, Annie LeBlanc, Alberta Health Services, Stephanie Hastings, Branden Manns, Lynora Saxinger

**Author notes:** Contact: Simon Décary, PT, PhD, Assistant Professor of Rehabilitation, Patient-Oriented Rehabilitation Lab (SPOR-REHAB) University of Sherbrooke, Faculty of Medicine and Health Sciences School of Rehabilitation, Research Centre of the CHUS, CIUSSS de l’Estrie-CHUS, 3001, 12e Avenue Nord, Sherbrooke, Quebec, Canada J1H 5N4. For and in close collaboration with: Alberta Health Services Stephanie Hastings, PhD Branden Manns, MD, MSc Lynora Saxinger, PhD. **Suggested citation:** Decary S, Dugas M, Stefan T, Langlois L, Skidmore B, Bhéreur A, and LeBlanc A. (2021). Care Models for Long COVID – A Rapid Systematic Review. SPOR Evidence Alliance, COVID-END Network. **For more information, please contact:** Simon Decary, PT, PhD.

## Abstract

**Context:** More than 18M people worldwide (150K Canadians) are living with Long COVID resulting in debilitating sequalae and disabilities that impact their quality of life and capacity to return to work. A new care model is needed for persons living with this complex and multi-systemic disease.

**Objectives:** What is the best-available evidence about care models for persons living with Long COVID?

**Design:** Rapid Living Systematic Review.

**Method:** We systematically searched seven electronic databases (MEDLINE, Embase, Web of Science, COVID-END, L-OVE, CDRS and WHO Ovid) on May 27^th^, 2021. Two independent reviewers screened titles, abstracts and full text. We included studies reporting on 1- persons living with Long COVID and 2- proposing a specific care model (i.e., dedicated clinic, care pathway). We extracted characteristic of studies (e.g., countries, study design, age group), referral pathways targeted (e.g., hospitalized, community), reporting of the care model implementation with number of patients, clinical settings of care model (e.g., primary care), healthcare professions included in the care model, care model principles (e.g., person-centred care) and care model components (e.g., standardized symptoms assessment). We used descriptive statistics and frequency count.

**Results:** We screened 2181 citations, read 65 full text and included 12 eligible articles reporting on care models for Long COVID. Half studies were from the United Kingdom. 7 out of 12 models reported conceptual models without a description of implementation. All but one model was designed for discharge and long-term follow-up of hospitalized patients and half models were designed for non-hospitalized or patients who lived with the disease only in the community. Nine out of 12 care models included primary care, 8 out of 12 included specialized clinics and all studies included rehabilitation services. A total of 30 healthcare professions and medical specialties were proposed for staffing Long COVID services. More than half studies proposed multidisciplinary teams, integrated/coordination of care, evidence-based care and patient-centred care as key care model principles. Standardized symptom assessment, follow-up system and virtual care were the most frequent care model components.

**Conclusion:** The implementation of care models for Long COVID is underway in several countries. Care models need to include both hospitalized and non-hospitalized patients. A complete care model for this population appears to design a care pathway integrating primary care, rehabilitation services and specialized clinics for medical assessment. The entry into care pathways is likely possible through a centralized referral system. It is possible to design sustainable and equitable care pathways for Long COVID in Canada integrated in current infrastructure.

**Protocol/Topic Registration:** CRD42021282266

**Summary:** An estimated 150K Canadians, mostly women, are facing debilitating sequalae and disabilities from Long COVID that impact their quality of life and capacity to return to work. A new care model is needed for persons with this complex and multi-systemic disease. We identified international care models describing the integration of primary care, rehabilitation services and specialized assessment clinics for Long COVID.

**Implications:** Limited evidence from this review of international care models for Long COVID point out to a care model for the Canadian context that should be co- designed with patients, clinicians, decision makers and researchers, and include: 1- A coordination unit to centrally receive referrals from both hospitalized and community-based patients; 2- Training of primary care teams to screen and support medical needs; 3- Integrated local multidisciplinary rehabilitation services; and 4- Access to medical specialty clinics for advanced testing and diagnoses.

**What is the current situation?:**

- More than 150K Canadians are with living the affliction of Long COVID, the patient-led term to describe long-term consequences of COVID-19. Long COVID is a multi-systemic and unpredictable disease impacting quality of life and return to work in middle aged population. To avoid widespread long-term disabilities impacting public health, Canadian provinces are seeking to organize a sustainable and equitable care model for Long COVID.

**What is the objective?:**

- To provide the best-available evidence about care models for persons living with Long COVID.

**How was the review conducted?:**

- We systematically searched seven electronic databases (MEDLINE, Embase, Web of Science, COVID-END, L-OVE, CDRS and WHO Ovid) on May 27th, 2021.
- Two independent reviewers screened title, abstract and full text.
- We included studies reporting on 1- persons living with Long COVID (post- hospitalized and community based) and 2- a specific care model (i.e., dedicated clinic, care pathway).
- We extracted characteristic of studies, referral pathways, clinical settings of care model, healthcare professions included in the care models, care model principles, care model components and reporting of the care model implementation.

**What did the review find?:**

- We found 12 international care models for Long COVID that covers follow-up of patients discharged following a hospitalization and patients who had lived the infection in the community.
- Most reported elements included in these care models were a coordination unit, primary care pathways, access to multidisciplinary rehabilitation and specialized medical services.
- The impact and costs of these care models are not yet reported.

## Context

Even if the pandemic ended today, more than 18M people worldwide (150K Canadians) would still be living the affliction of Long COVID, the patient-led term to describe long-term consequences of COVID-19. Long COVID translates into symptoms that develop during or following an infection from COVID-19 and continue for 4 weeks or more ^1, 2^. Update from the Office of National Statistics in the United Kingdom on April 1^st^, 2021, shows that 13.7% of all cases will exhibit symptoms longer than 12 weeks ^3^. The extensive online survey of Patient-Led Research for COVID-19 marked a turning point: 3,762 people living with Long COVID described more than 50 symptoms over 10 body systems that persisted over 6 months ^4^. All patients, hospitalized or not, are at risk of developing debilitating sequelae and many continue to experience complex symptoms 12 months after the infection with uncertain prognosis ^4–6^.

In Canada, specialized post-COVID clinics are a rare occurrence. Primary care is in a precarious state and regions lack knowledge and organizational readiness. Physicians and nurses are often untrained to diagnose and refer patients to appropriate services. Community-based physical and occupational therapists are frequently unaware of safety issues regarding rehabilitation after COVID-19. On June 10^th^, 2021, the CIHR Knowledge Mobilization Forum “Moving Forward Together: Recognizing Lived Experience, Optimizing Appropriate Rehabilitation and Enabling Research on Post COVID-19 Condition (‘Long COVID’) in Canada” taught us the experience of care from persons living with Long COVID: stigmatization by clinicians, financial insecurity, lack of recognition by insurances and fearful prognosis. People with Long COVID need patient-centered and evidence-based care to prevent long-term disabilities. A new care model for the Canadian context is needed for persons with this complex disease ^7^.

## Research question

What is the best-available evidence about care models for persons living with Long COVID?

## Methods

We performed a rapid systematic review following the Joanna Briggs Institute’s Manual for Evidence Synthesis ^8^ and report our findings according to the PRISMA guidelines ^9^.

### Literature Search

We systematically searched seven databases: MEDLINE, Embase, Web of Science, COVID-END, L- OVE, CDSR, and WHO Ovid. An experienced medical information specialist (Skidmore) developed and tested the search strategies through an iterative process in consultation with the review team. Using the multifile option as well as the deduping tool in Ovid, we searched Ovid MEDLINE®, including Epub Ahead of Print, In-Process & Other Non-Indexed Citations, and Embase Classic+Embase. We also searched Web of Science (Core Collection) and the Cochrane Database of Systematic Reviews. We completed a rapid grey literature search by scanning key healthcare organizations’ websites (ex: NHS) to identify unpublished care models for Long Covid. We executed all searches on May 27^th^, 2021.

The search strategy included terms related to 1- Long COVID and its currently known variation (e.g., post-Covid syndrome) and 2- Care models (e.g., clinics model, pathways). We used a mixture of controlled vocabulary (e.g., “COVID-19”, “Long Term Adverse Effects”, “Recovery of Function”) and free-text terms (e.g., “long COVID”, “longhaul”, “chronic symptoms”) and applied filters for care models. We did not apply any date or language restrictions. See Appendix 3 for a copy of the search strategies as executed. We downloaded and deduped the records using EndNote 9.3.3 (Clarivate) and uploaded to DistillerSR (Evidence Partners).

### Eligibility Criteria

Our inclusion criteria (PICO) were as followed:

|  |  |
| --- | --- |
| **Population:** | Persons living with Long COVID (children and adults) or healthcare professionals managing Long COVID. We included all definition of persistent symptoms of COVID-19 (e.g., post-COVID syndrome). |
| **Intervention:** | Care models (including pathways, trajectories, frameworks or structured clinics) to organise health care services for Long COVID. |
| **Comparator:** | Natural recovery, usual care models, inter-jurisdiction models comparison if available. |
| **Outcomes:** | Any outcomes or specifically system-level outcomes (e.g., cost, access, quadruple aim), patient-level outcomes (e.g., specific PROMS related to symptoms), clinician-level outcome (e.g., satisfaction with the care pathways). |
| **Study design:** | Any design. |
| **Settings:** | Any setting. |

### Study Selection

We developed and pilot-test standardized forms for titles & abstracts and full text screening. Study selection was conducted by pairs of two reviewers independently. Discrepancies were resolved by discussion or by a third senior reviewer. We conducted a pilot exercise with 10 citations for each phase.

### Data Extraction

We developed a standardized form for data extraction, which we piloted on 3 articles. All data was extracted by one of two reviewers and a third reviewer verified all extracted data. Discrepancies was resolved by discussion or by a third senior reviewer. We extracted characteristic of studies (e.g., countries, study design, age group), targeted referral pathways (e.g., hospitalized, community), reporting of the care model implementation with number of patients, clinical settings of care model (e.g., primary care), characteristics of the care model (ex: funding, staffing, etc.), healthcare professions included in the care models, care model principles (e.g., person-centered care), care model components (e.g., standardized symptoms assessment) and any collected outcomes.

### Risk of Bias Assessment

As a rapid review, we identified and described best evidence of care models. We did not completed risk of bias assessment since most studies were conceptual papers.

### Data Synthesis

We analyzed the extracted data using a traditional content analysis and performed a thematic analysis to classify emerging themes regarding best practices surrounding care models (see definition in Appendix 2). We present our results in a narrative format.

### Stakeholders’ Engagement

Three knowledge users from the Alberta Health Services actively participated in this rapid review. They were involved in defining the research topic and objectives. They received a weekly update via email communications. We engaged one patient-partner who helped reviewed the PICO, data extraction, reporting and read all final material.

## Results

### Literature Search

We identified 2184 citations, screened 1806 titles and abstract, 65 full text and included 12 articles reporting on care models for Long COVID (Figure 1) ^10–21^.

**Fig. 1.**
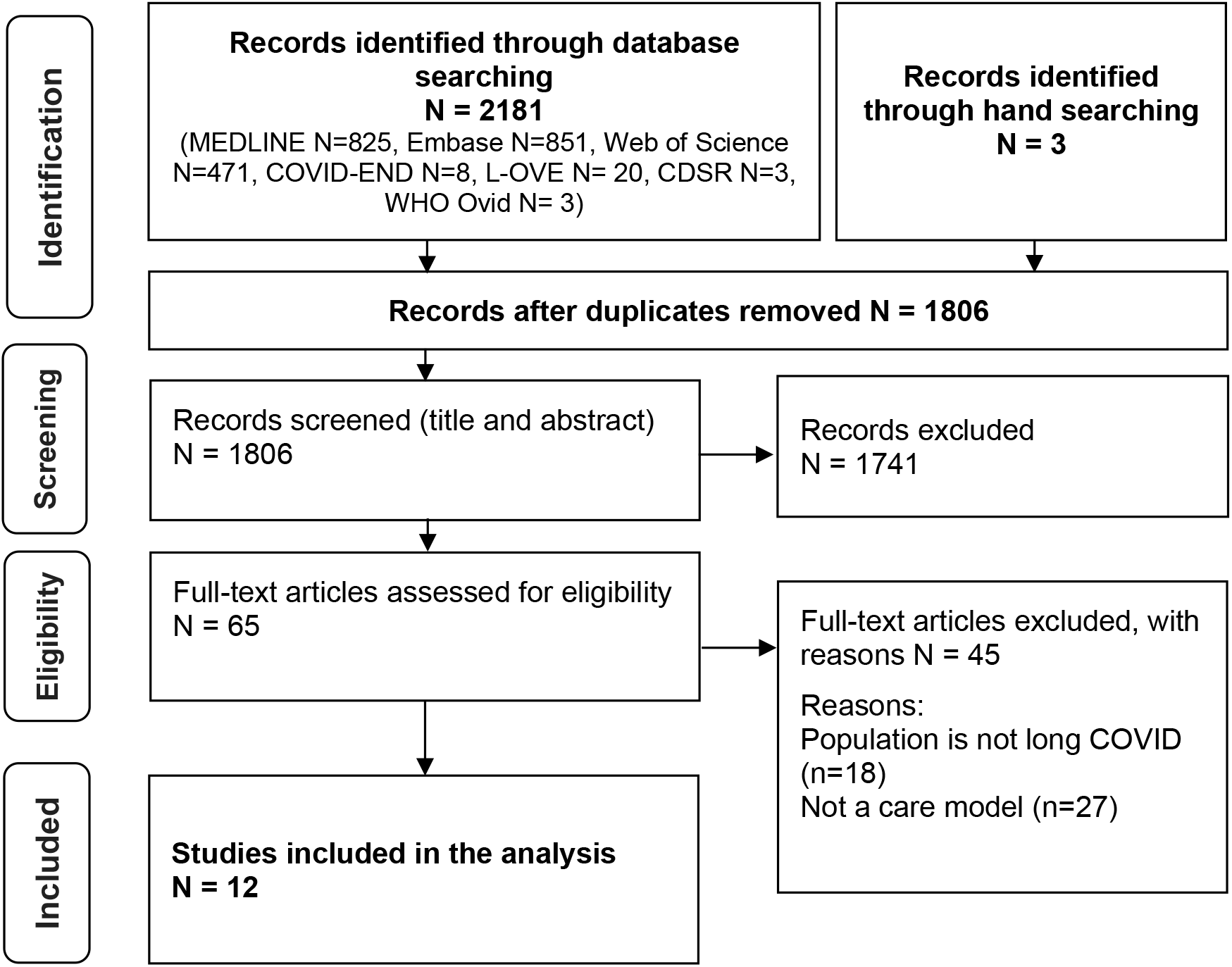
PRISMA Flow Diagram.

### Characteristics of Included Studies

Of the 12 included studies (Table 1), six were from the United Kingdom and three from the United States. Seven were descriptive studies of care models concepts, two were literature review with a proposed model, one was a qualitative study, one a was a cross-sectional survey and one study reported a service evaluation of a proposed post-Covid clinic. All studies reported on adult population and one model included children. All studies covered care models for post-hospitalized patients and six studies further reported on care models for patients in the community. Seven studies reported having implemented their care model in clinical settings. All studies included rehabilitation services, nine integrated primary care and eight had a specialty care component.

**Table 1.**
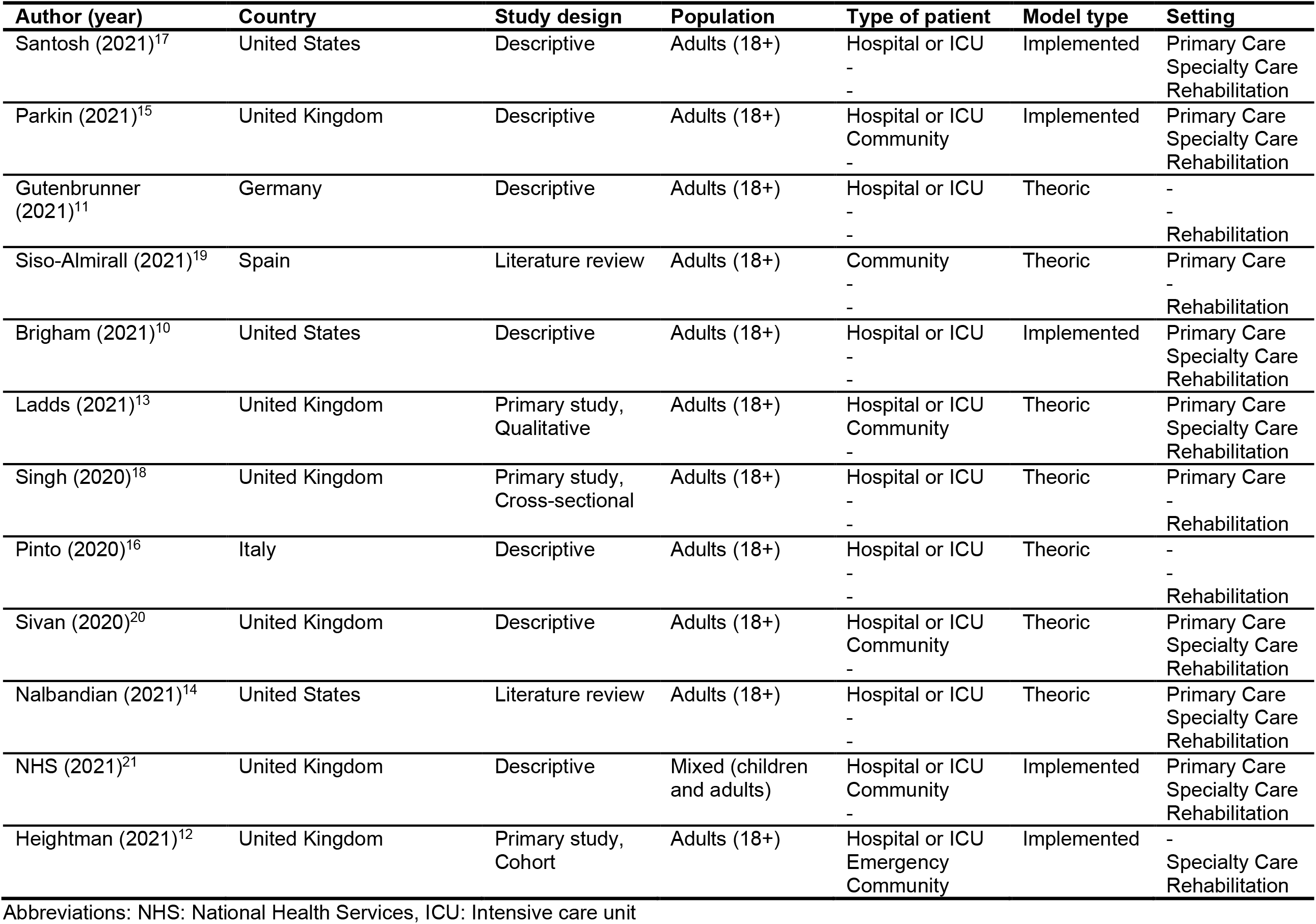
Study Characteristics.

| Author (year) | Country | Study design | Population | Type of patient | Model type | Setting |
| --- | --- | --- | --- | --- | --- | --- |
| Santosh (2021)<sup>17</sup> | United States | Descriptive | Adults (18+) | Hospital or ICU<br>-<br>- | Implemented | Primary Care<br>Specialty Care<br>Rehabilitation |
| Parkin (2021)<sup>15</sup> | United Kingdom | Descriptive | Adults (18+) | Hospital or ICU<br>Community<br>- | Implemented | Primary Care<br>Specialty Care<br>Rehabilitation |
| Gutenbrunner (2021)<sup>11</sup> | Germany | Descriptive | Adults (18+) | Hospital or ICU<br>-<br>- | Theoric | -<br>-<br>Rehabilitation |
| Siso-Almirall (2021)<sup>19</sup> | Spain | Literature review | Adults (18+) | Community<br>-<br>- | Theoric | Primary Care<br>-<br>Rehabilitation |
| Brigham (2021)<sup>10</sup> | United States | Descriptive | Adults (18+) | Hospital or ICU<br>-<br>- | Implemented | Primary Care<br>Specialty Care<br>Rehabilitation |
| Ladds (2021)<sup>13</sup> | United Kingdom | Primary study, Qualitative | Adults (18+) | Hospital or ICU<br>Community<br>- | Theoric | Primary Care<br>Specialty Care<br>Rehabilitation |
| Singh (2020)<sup>18</sup> | United Kingdom | Primary study, Cross-sectional | Adults (18+) | Hospital or ICU<br>-<br>- | Theoric | Primary Care<br>-<br>Rehabilitation |
| Pinto (2020)<sup>16</sup> | Italy | Descriptive | Adults (18+) | Hospital or ICU<br>-<br>- | Theoric | -<br>-<br>Rehabilitation |
| Sivan (2020)<sup>20</sup> | United Kingdom | Descriptive | Adults (18+) | Hospital or ICU<br>Community<br>- | Theoric | Primary Care<br>Specialty Care<br>Rehabilitation |
| Nalbandian (2021)<sup>14</sup> | United States | Literature review | Adults (18+) | Hospital or ICU<br>-<br>- | Theoric | Primary Care<br>Specialty Care<br>Rehabilitation |
| NHS (2021)<sup>21</sup> | United Kingdom | Descriptive | Mixed (children and adults) | Hospital or ICU<br>Community<br>- | Implemented | Primary Care<br>Specialty Care<br>Rehabilitation |
| Heightman (2021)<sup>12</sup> | United Kingdom | Primary study, Cohort | Adults (18+) | Hospital or ICU<br>Emergency<br>Community | Implemented | -<br>Specialty Care<br>Rehabilitation |

Abbreviations: NHS: National Health Services, ICU: Intensive care unit

### Care Models Principles

We identified a total of 22 different principles (Table 2). The five most common principles included multidisciplinary teams (92%), integrated care (67%), self-management (58%), coordination of care (58%) and evidence-based care (58%).

**Table 2.**
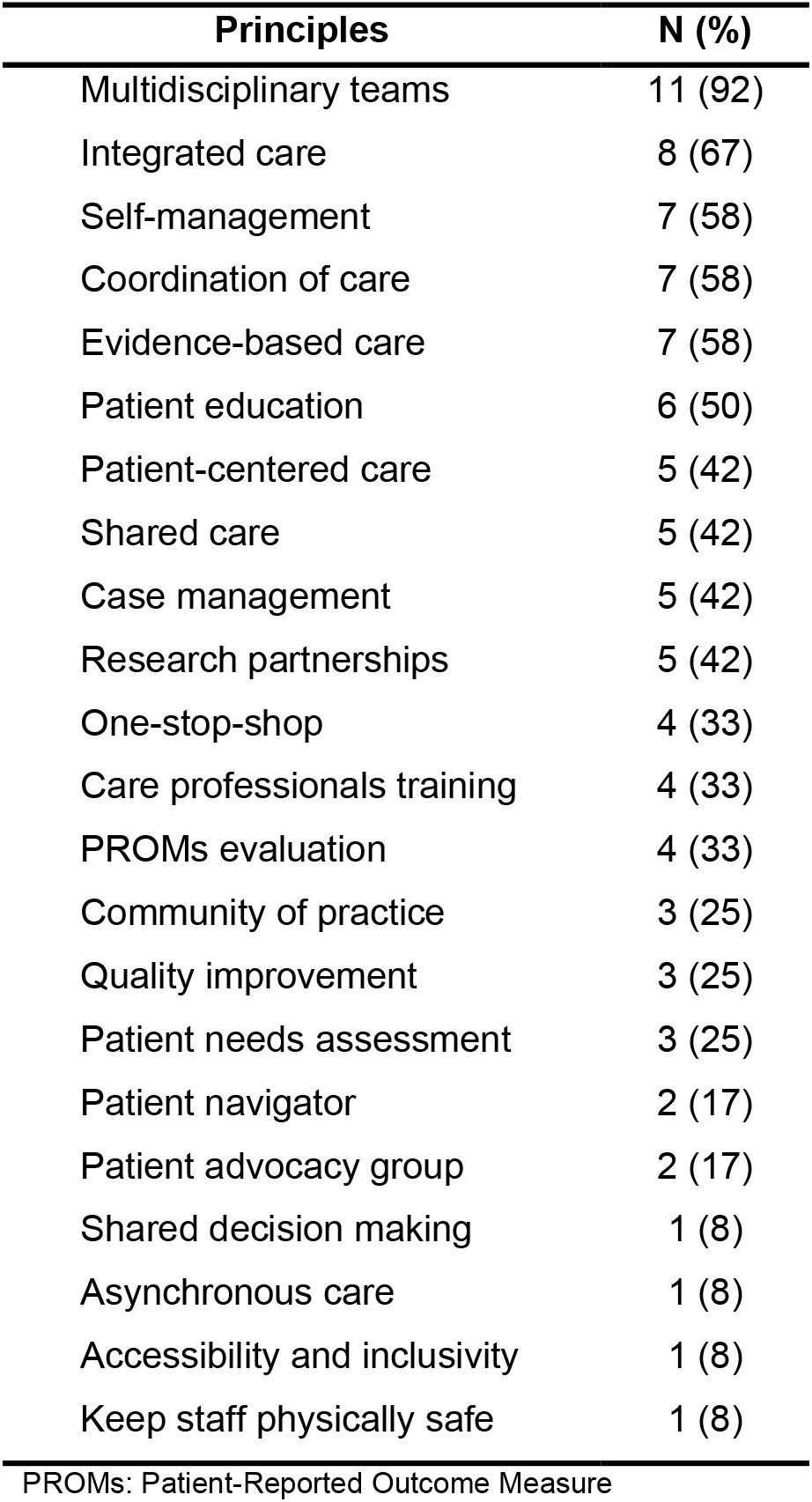
Care Model Principles.

| Principles | N (%) |
| --- | --- |
| Multidisciplinary teams | 11 (92) |
| Integrated care | 8 (67) |
| Self-management | 7 (58) |
| Coordination of care | 7 (58) |
| Evidence-based care | 7 (58) |
| Patient education | 6 (50) |
| Patient-centered care | 5 (42) |
| Shared care | 5 (42) |
| Case management | 5 (42) |
| Research partnerships | 5 (42) |
| One-stop-shop | 4 (33) |
| Care professionals training | 4 (33) |
| PROMs evaluation | 4 (33) |
| Community of practice | 3 (25) |
| Quality improvement | 3 (25) |
| Patient needs assessment | 3 (25) |
| Patient navigator | 2 (17) |
| Patient advocacy group | 2 (17) |
| Shared decision making | 1 (8) |
| Asynchronous care | 1 (8) |
| Accessibility and inclusivity | 1 (8) |
| Keep staff physically safe | 1 (8) |

PROMs: Patient-Reported Outcome Measure

### Care Models Components

We identified a total of 10 distinct care model components (Table 3). The five most common components included standardized symptom assessment (92%), referral system (83%), follow-up system (83%), virtual care (83%), and home-base care (58%).

**Table 3.**
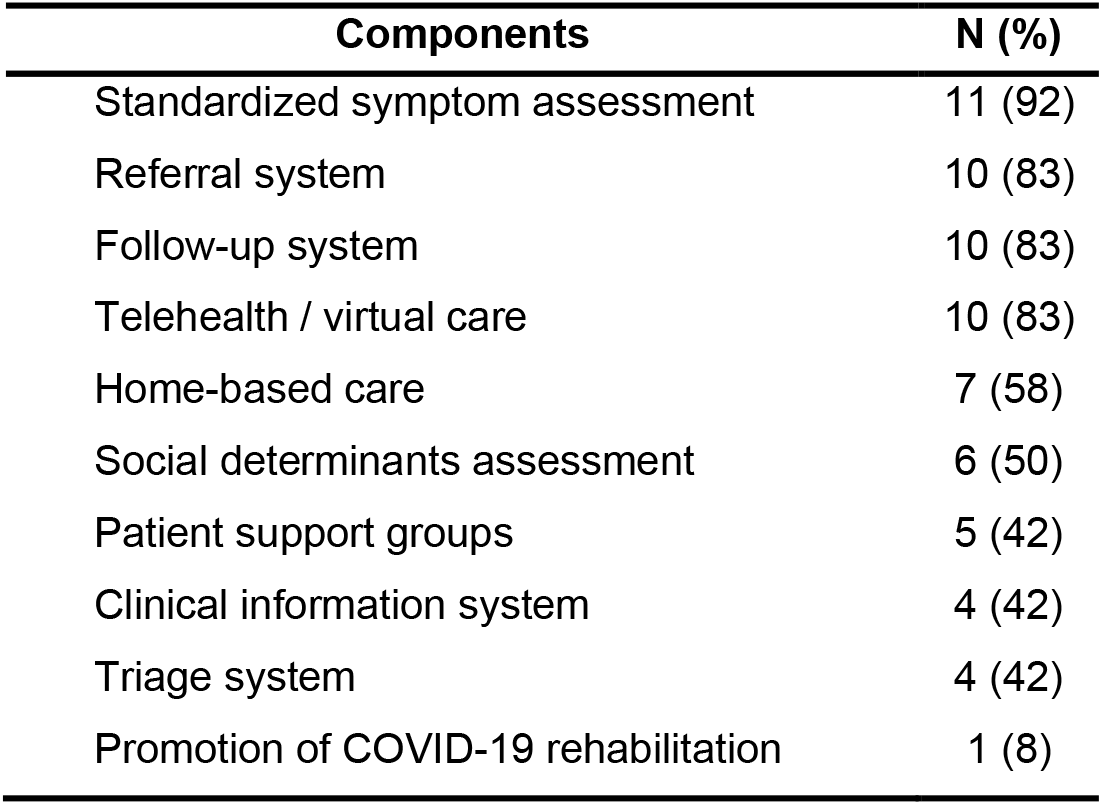
Care models components.

### Healthcare Professionals and Medical Specialties Included in Care Models

We identified all healthcare professionals and medical specialties included in care models (Table 4). The ten professionals most embedded in these models included pulmonary/respiratory specialists (100%), cardiovascular specialists (92%), psychiatry and psychology (83%), physiotherapy (83%), occupational therapy (75%), social work (75%), neurology specialists (75%), primary care physicians (58%), nutrition (58%) and speech and language therapists (50%).

**Table 4.**
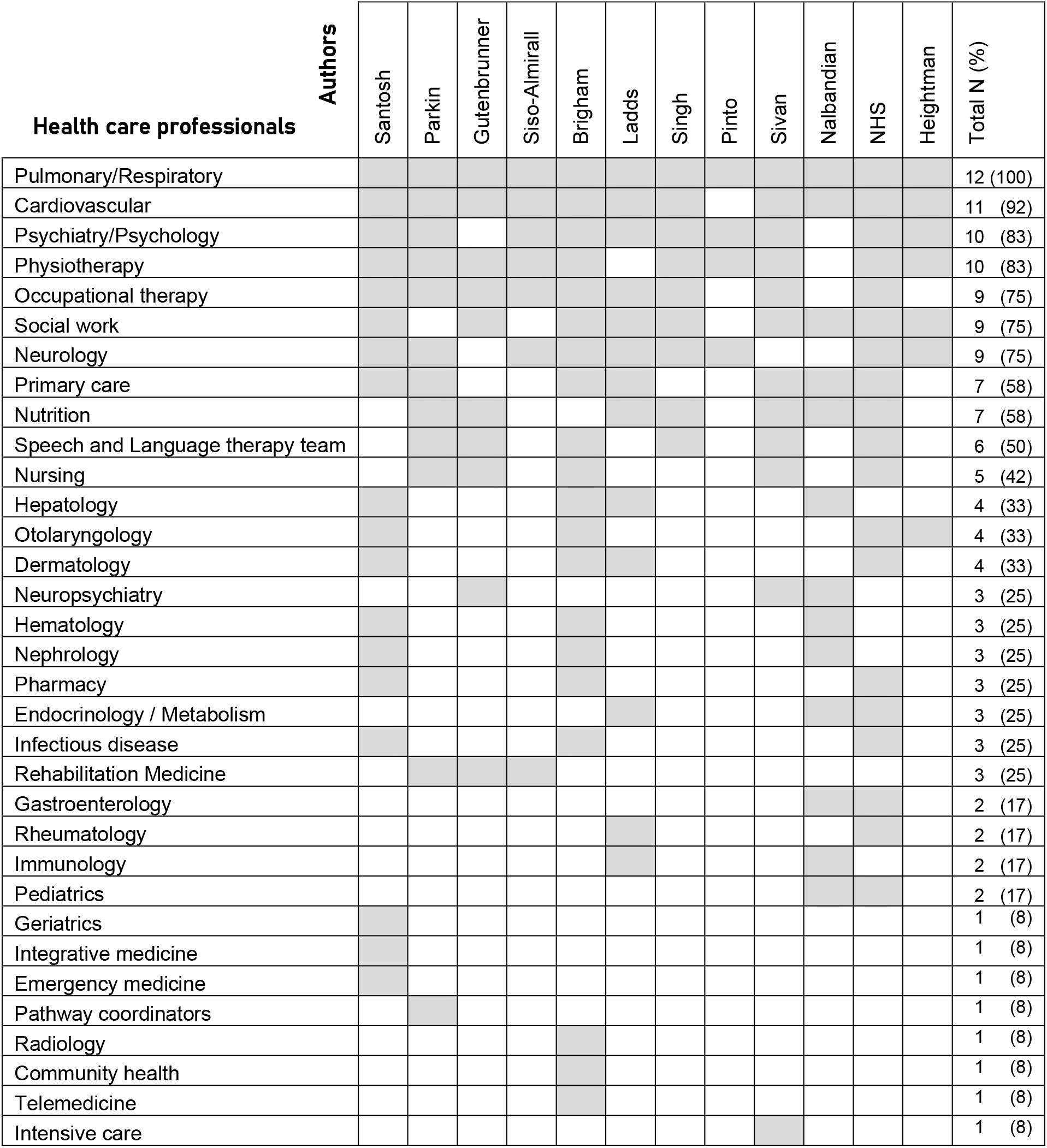
Healthcare Professionals and Medical Specialties Included in Care Models.

### Impact

None of the included studies provided impact analysis. *Heightman et al.* conducted a service evaluation of the first 12-month experience of the first UK dedicated post-COVID clinic in the United Kingdom.

Their analysis is limited to healthcare utilization (e.g., diagnostic tests) of 1325 assessed individuals and showed similar rates of onward specialist referrals between non-hospitalized and hospitalized patients and incapacity of 50% of the cohort to return to work full-time at their first assessment.

### Costs

The “National guidance for post-COVID syndrome assessment clinics” from the United Kingdom is the only included reference to document a nation-wide investment in Long COVID care. The total investment to set-up 89 post-Covid clinics was £34 million (58.5M$CAN) between December 2020 and April 2021. A new guidance document (unreported at the time of our search strategy) providing cost to extend services in the United Kingdom model is explained below.

## Discussion

The objective of this rapid systematic review was to describe the best-available evidence about care models for persons living with Long COVID. We found 12 international care models that followed patients discharged following a hospitalization and patients who had lived the infection in the community. The most reported elements in these care models included coordination units, primary care pathways, access to multidisciplinary rehabilitation and specialized medical services. The impact and long-term costs of these care models are not yet established. Overall, evidence remains scarce.

Initial care models that emerged in the early months of the pandemic were centered on “post-COVID clinics” ^10, 13, 14, 22^. These clinics were vital in the ecosystem for Long COVID as they allow specialized assessment and act as a gateway for medical diagnoses. Unfortunately, shortcomings started appearing such as long waiting lists, difficulties training clinicians, barriers to access for patients with fatigue living outside city areas, absence of integrated multidisciplinary rehabilitation and long-term funding sustainability, warranting a more integrated care model for Long COVID. Critical and central to this proposed integrated care model was primary care ^13, 22–27^.

While we identified frequent care model principles and components, most were not thoughtfully described, limiting our understanding of the underlying structure to be put in place. Particularly, most model proposed a place for primary care for the long-term management of Long COVID without describing its execution. This further creates uncertainties around impact and cost, and how to appropriately implement and scale-up these care models. Future research will need to describe and demonstrate how each proposed principles and components are implemented and link to observed outcomes. Furthermore, only a small sample of studies reported estimated costs of these care models. This lack of information prevents from determining the impact and cost-effectiveness of implementing these models.

A wide diversity of healthcare professionals and medical specialties have been proposed to care for Long COVID. While we observed trends concerning the commonly proposed team members to care for Long COVID, identifying a more defined core will be essential to designing a care model that is efficient, feasible, and scalable. Indeed, some models presented advocated for the inclusion of numerous (>10) medical specialists and health professionals in the care of a patient, which with the current health setting and staff shortage may render such model unfeasible. Future research should aim to determine the exact implication of each proposed healthcare professions and the parameters of their implication in the care models (i.e., objective of the role, number of interventions).

Lastly, quality standards for patient-centered care were proposed throughout the identified models, highlighting the need for and importance of high-quality and personalized care. Unfortunately, these standards were not operationalized nor measured in the reported models. Identifying processes and outcomes that matter to patients suffering from Long COVID and are centered around their goals toward recovery will be critical.

## Limitations

As a rapid review, we did not search all available databases and grey literature and some existing care models may have been missed. The search strategy was not duplicated by a second information specialist and may have missed relevant articles. We did not conduct a systematic risk of bias appraisal as most included article reported only descriptive and conceptual underpinnings of care models for Long COVID.

## Contextualization

As June 18^th^, 2021, the province of Alberta had 231K cases of COVID-19. Given conservative estimates that 13.7% of cases will continue to experience persistent symptoms impacting quality of life for at least 12 weeks, the Alberta Health System could face a surge of 32K cases of Long COVID. Many of these patients may require care for at least one year.

Unfortunately, at this point in time, the evidence regarding best practices and effectiveness of care models for Long COVID remains scarce. Based on our limited findings and the current context that warrants immediate action, the Alberta Health Services could invest in the co-design with patients, clinicians, decision makers and researchers of a care model for Long COVID integrated in its current infrastructure. The care model could include four elements: 1- A coordination unit to centrally receive referrals from both hospitalized and community-based patients (e.g., leverage AHS’s centralized phone infrastructure with proposed standardized assessment instead of dedicated new clinics as access points (UK model)), 2- Training of primary care teams to screen and support medical needs, 3- Integrated local organization of multidisciplinary rehabilitation teams and 4- Access to medical specialty clinics for advanced testing and diagnoses. These four elements can work as a network putting the needs of each person living with Long COVID at the center. The Infographic (next page) schematize this possible care model. As for cost, the United Kingdom announced a total investment of 230M$CAN to care for an estimated 1.1M Long COVID cases. At scale for Alberta, projections could require a 6.7M$ investment for 2021-2022 fiscal year.

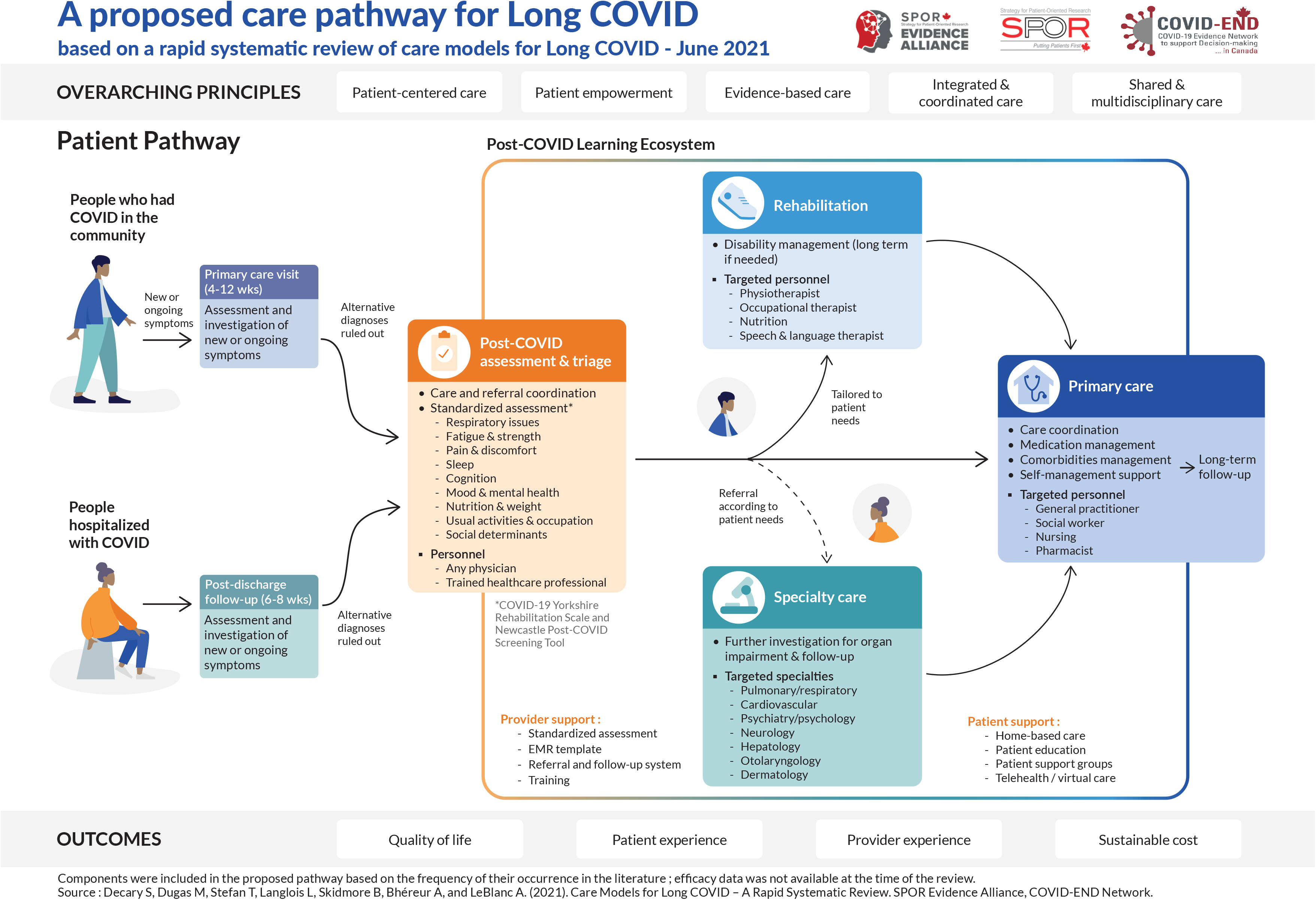

## Data Availability

All data produced in the present study are available upon reasonable request to the authors

## Funding Acknowledgements

This review was funded by the Alberta Health Services, the SPOR Evidence Alliance ([SPOR EA](http://www.sporevidencealliance.ca/)), and the COVID-19 Evidence Network to support Decision-making (COVID-END).

The SPOR Evidence Alliance ([SPOR EA](http://www.sporevidencealliance.ca/)) is supported by the Canadian Institutes of Health Research ([CIHR](http://www.cihr-irsc.gc.ca/e/193.html)) under the Strategy for Patient-Oriented Research ([SPOR](http://www.cihr-irsc.gc.ca/e/41204.html)) initiative.

COVID-19 Evidence Network to support Decision-making (COVID-END) is supported by the Canadian Institutes of Health Research ([CIHR](http://www.cihr-irsc.gc.ca/e/193.html)) through the Canadian 2019 Novel Coronavirus (COVID-19) Rapid Research Funding opportunity.

## Third-Party Materials

If you wish to reuse non-textual material from this report that is attributed to a third party, such as tables, figures or images, it is your responsibility to determine whether permission is required for such use and to obtain necessary permission from the copyright holder. The risk of claims resulting from infringement of any third-party-owned material rests solely with the user.

## General Disclaimer

This report was prepared by Simon Décary’s and Annie LeBlanc’s team on behalf of the SPOR Evidence Alliance and COVID-END. It was developed through the analysis, interpretation and synthesis of scientific research and/or health technology assessments published in peer-reviewed journals, institutional websites and other distribution channels. It also incorporates selected information provided by experts and patient/citizen partners with lived experience on the subject matter. This document may not fully reflect all the scientific evidence available at the time this report was prepared. Other relevant scientific findings may have been reported since completion of this synthesis report.

SPOR Evidence Alliance, COVID-END and the project team make no warranty, express or implied, nor assume any legal liability or responsibility for the accuracy, completeness, or usefulness of any information, data, product, or process disclosed in this report. Conclusions drawn from, or actions undertaken on the basis of, information included in this report are the sole responsibility of the user.

## APPENDICES

## APPENDIX 1 Inspiring Practices Showcase

Our review presents a descriptive overview of international care models for Long COVID. Here we provide more detailed vignettes of key care models specifically for primary care, rehabilitation services and post-discharge speciality clinics.

### The NHS Post-COVID-19 Clinics

**Country**: United Kingdom.

**Description**: Nationwide integrated care pathways that include post-COVID-19 assessment clinics.

**Population**: Adults & children, ongoing and post-COVID-19, for community and post-discharged patients.

**Model Principles**: Patient-centered care, accessibility and equity, integrated and shared care, continuity of care, one-stop-shop, multidisciplinary teams.

**Model Components**: Standardized holistic symptoms assessment, patient needs assessment, triage, referral and follow-up system, shared decision making, self-management, patient education, patient support and advocacy groups, training for healthcare professionals, PROMs/PREMs evaluation, clinical information system, telehealth and virtual care, research partnership.

**Setting**: Primary care, speciality care, rehabilitation.

**Funding**: £10 million (October 2020), £24 million (March 2021 to 2022), £50 million for research.

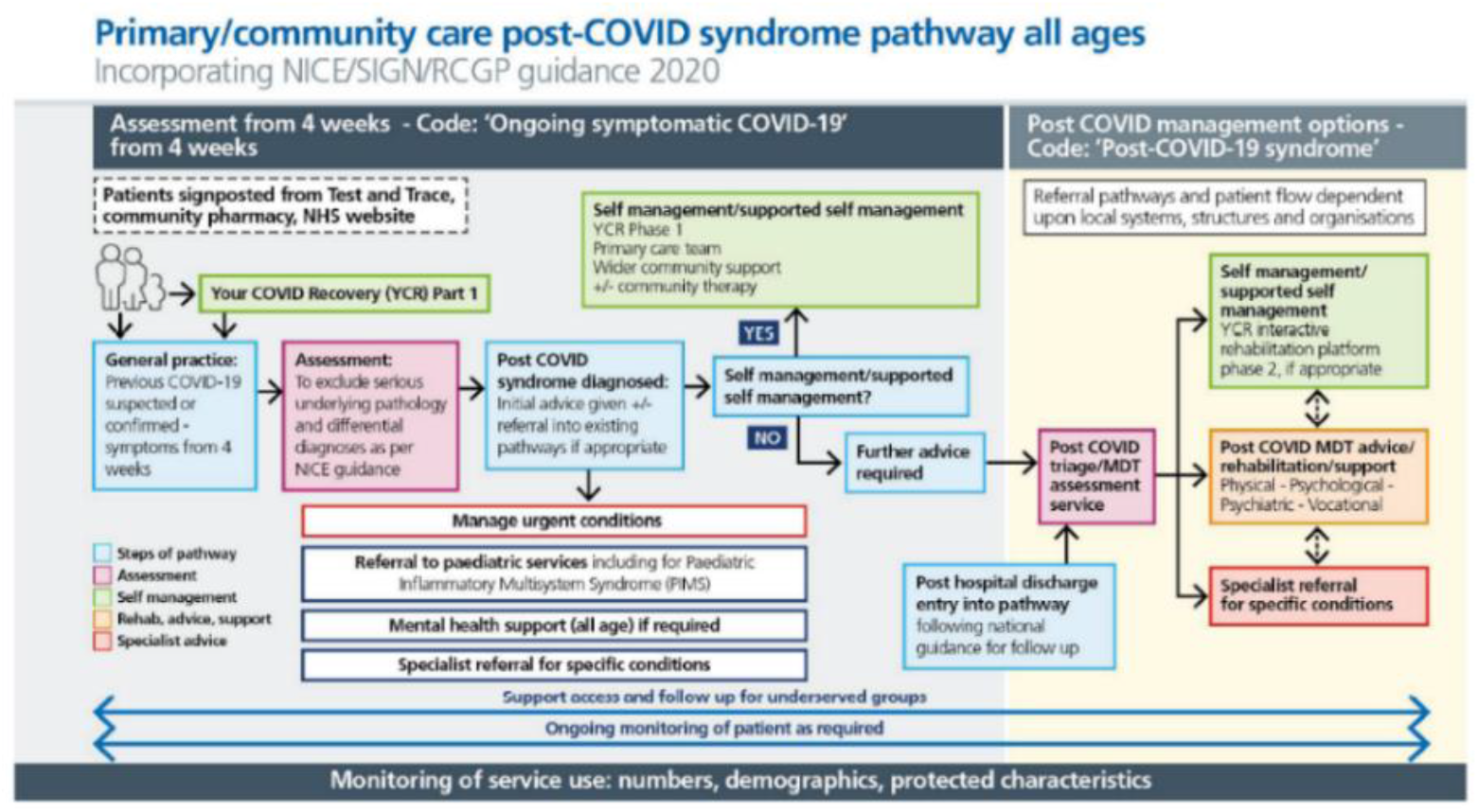

Reference: National Health Service (NHS). National guidance for post-COVID syndrome assessment clinics, Version 2, 26 April 2021. Available from https://www.england.nhs.uk/coronavirus/wp-content/uploads/sites/52/2020/11/C1248-national-guidance-post-covid-syndrome-assessment-clinics-v2.pdf

### The PACT / OPTIMAL Post-COVID-19 Clinics

**Country**: United States.

**Description**: Multidisciplinary post-COVID-19 clinics at two academic health systems, Johns Hopkins and the University of California-San Francisco.

**Population**: Adults, ongoing and post-COVID-19, for community and post-discharged patients.

**Model Principles**: Patient-centered care, integrated and shared care, multidisciplinary teams, continuity of care, one-stop-shop, asynchronous care, evidence-based care, quality improvement, research partnership.

**Model Components**: Standardized holistic symptoms assessment, referral and follow-up system, patient needs assessment, case management, patient navigator, community of practice, PROMs evaluation, training for healthcare professionals, clinical information system, home-based care, telehealth and virtual care.

**Setting**: Primary care, speciality care, rehabilitation.

**Costs**: Cost-neutral for initial implementation, currently seeking funding for staff and additional physical space.

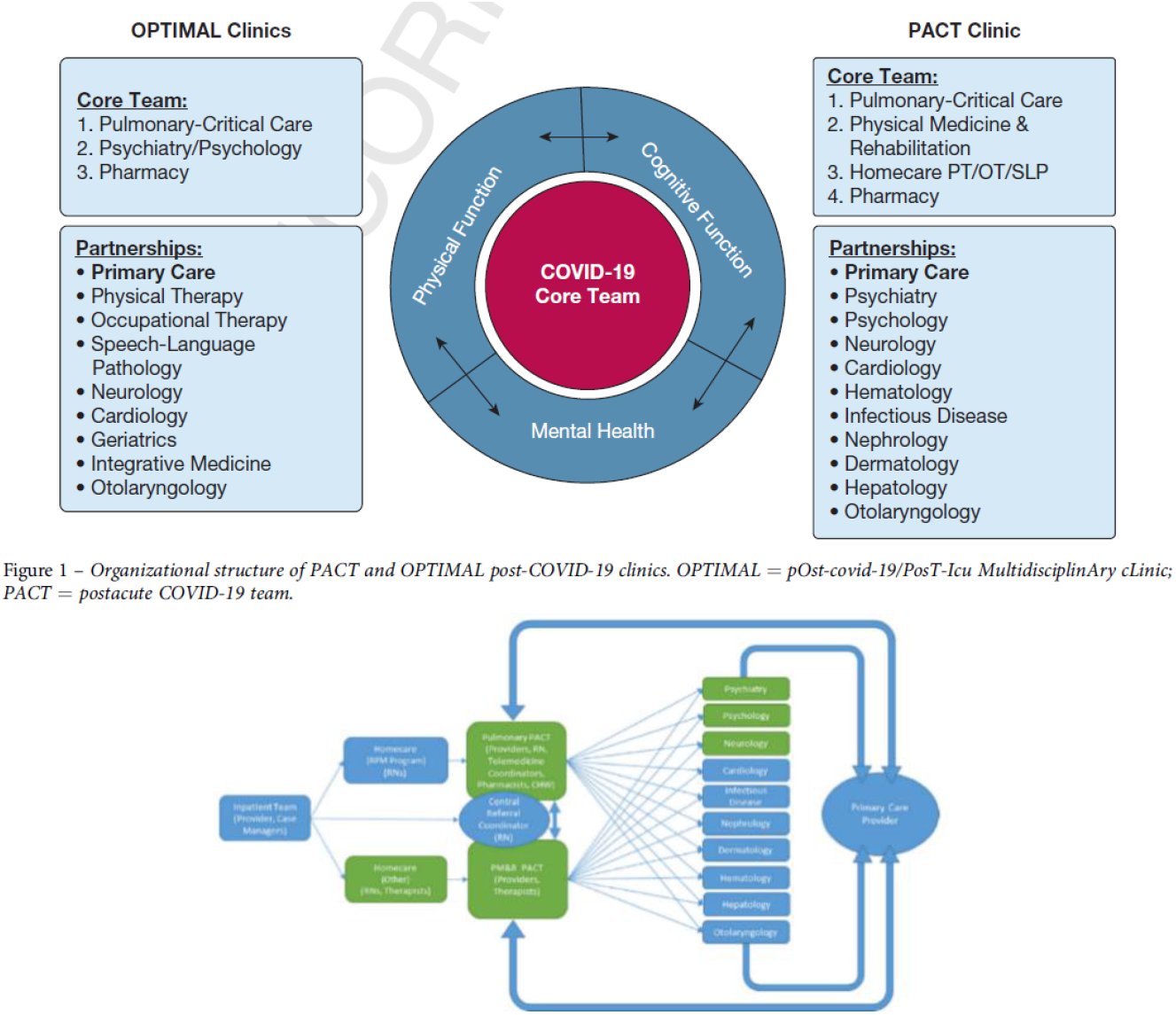

References: Santhosh L, Block B, Kim SY, Raju S, Shah RJ, Thakur N, et al. How I Do It: Rapid Design and Implementation of Post-COVID-19 Clinics. Chest. 2021;31:31. And : Brigham E, O’Toole J, Kim SY, Friedman M, Daly L, Kaplin A, et al. The Johns Hopkins Post-Acute COVID-19 Team (PACT): A Multidisciplinary, Collaborative, Ambulatory Framework Supporting COVID-19 Survivors. Am J Med. 2021;134:462-7.e1.

### The Leeds Rehabilitation Pathway

**Country**: United Kingdom.

**Description**: Community rehabilitation pathway using a telephone screening tool and a multidisciplinary team for complex case management.

**Population**: Adults, ongoing and post-COVID-19, for community and post-discharged patients.

**Model Principles**: Integrated and shared care, continuity of care, multidisciplinary teams.

**Model Components**: Standardized holistic symptoms assessment, referral and follow-up system, patient education, self-management, case management, telehealth and virtual care.

**Setting**: Primary care, speciality care, rehabilitation.

**Costs**: Not reported.

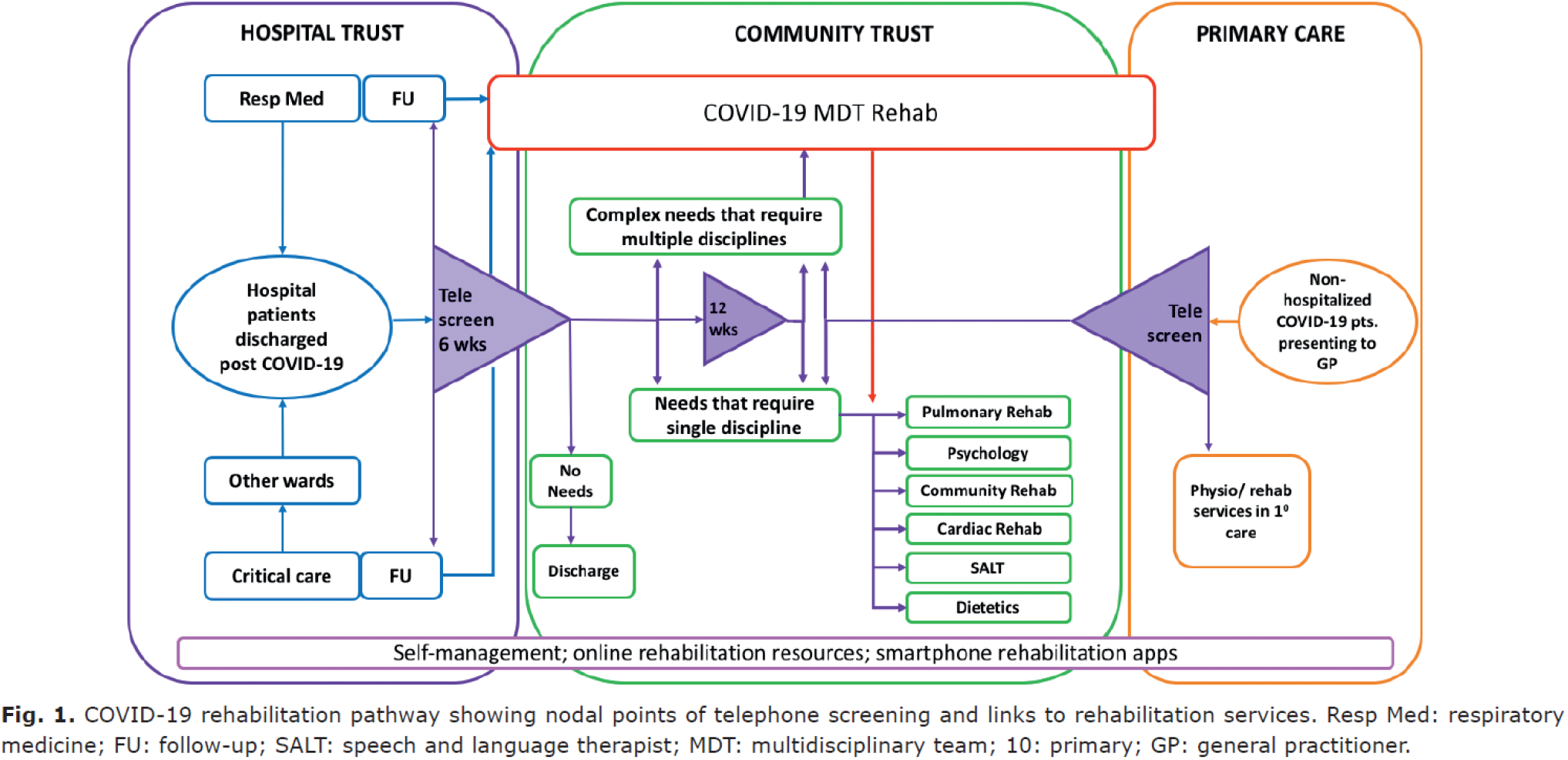

Reference: Sivan M, Halpin S, Hollingworth L, Snook N, Hickman K, Clifton IJ. Development of an integrated rehabilitation pathway for individuals recovering from COVID-19 in the community. J Rehabil Med. 2020;52:jrm00089.

## APPENDIX 2 Definitions

### Care model principles

#### Patient-centered care

“Providing care that is respectful of, and responsive to, individual patient preferences, needs and values, and ensuring that patient values guide all clinical decisions.”32

#### Shared decision making

“An approach where clinicians and patients share the best available evidence when faced with the task of making decisions, and where patients are supported to consider options, to achieve informed preferences.”33

#### Patient education

“The process of influencing patient behavior and producing the changes in knowledge, attitudes and skills necessary to maintain or improve health.”34

#### Self-management

“Self-management support is the help given to people with chronic conditions that enables them to manage their health on a day-to-day basis.”35

#### Integrated care

“Health services that are managed and delivered so that people receive a continuum of health promotion, disease prevention, diagnosis, treatment, disease-management, rehabilitation and palliative care services, coordinated across the different levels and sites of care within and beyond the health sector, and according to their needs throughout the life course.”36

#### Multidisciplinary teams

“A multidisciplinary team involves a range of health professionals, from one or more organisations, working together to deliver comprehensive patient care.”37

#### Shared care

“The joint participation of primary care physicians and specialist care physicians in the planned delivery of care for patients with a chronic condition, informed by an enhanced information exchange over and above routine discharge and referral.”38

#### Continuity or coordination of care

“The delivery of a ’seamless service’ through integration, coordination and the sharing of information between different providers.”39

#### Case management

“The term “case manager” is generally used to describe a care professional that promotes upstream approaches to health care and facilitates individualized care coordination for those with complex and chronic health conditions. (…) Case managers are often known to conduct patient assessments; patient identification and outreach; care planning and coordination; and service evaluation.”40

#### Patient navigator

“A partnership between a patient or caregiver and a navigator (e.g. registered nurse or peer) that seeks to proactively guide patients through the healthcare continuum to facilitate timely access to care and foster self-management and autonomy through education and emotional support. (…) Patient navigation typically employs an individualized, holistic approach to help patients navigate through a range of health care services”40

#### One-stop-shop

This means that the patient sees all care team members and subspecialists in the same clinical visit.

#### Asynchronous care

Type of telehealth care that use “store-and-forward technologies to collect health data that can be transmitted and interpretated later”41. For example, the use of technology to remotely monitor patient outcomes with mobile apps.

#### Evidence-based care

“An approach to decision making where a clinician uses the best evidence available, in consultation with the patient (evidence-based patient choice), to decide upon which option best suits the patient.”42

#### Community of practice

“Groups of people who share a concern, a set of problems, and a passion about a topic, and who deepen their knowledge and expertise in this area by interacting on an ongoing basis.”43

#### Quality improvement

“The combined and unceasing efforts of everyone—healthcare professionals, patients and their families, researchers, payers, planners and educators—to make the changes that will lead to better patient outcomes (health), better system performance (care) and better professional development.”44

#### PROMs evaluation

Patient-reported outcome measures (PROMs). “PROMs are measurement tools that patients use to provide information on aspects of their health status that are relevant to their quality of life, including symptoms, functionality, and physical, mental and social health.”45

#### Training for healthcare professionals

Training for healthcare professionals that is planned in the care model (ex: information about long covid symptoms, assessment tools, referral pathways, etc.)

#### Research partnerships

Incorporating research aspects into the care model to study long covid or to evaluate the impact of the care model for long covid.

### Care model components

#### Decision support for healthcare professionals

The care model reports the use of decision supports for healthcare professionals. These include, for example, clinical decision support systems or decision trees. “Clinical decision support systems (CDSS) are computer-based programs that analyze data within EHRs to provide prompts and reminders to assist health care providers in implementing evidence-based clinical guidelines at the point of care.”46

#### Clinical information system

The care model reports the use or adaptation of a clinical information system to facilitate the management of Long Covid. For example, EMR adaptations, including forms specific for Long Covid, etc.

#### Triage system

Process by which patients are assessed and prioritized based on specific criteria.

#### Standardized symptoms assessment

Using standardized tools to assess patient’s symptoms.

#### Social determinants assessment

A step in the care model that involves assessing the patient’s psychosocial needs, such as employment/return to work, food security, housing situation, etc.

#### Referral system

Process by which patients are directed to other healthcare professionals to receive other type of care or tests (ex: referral to speciality care).

#### Follow-up system

Process by which a healthcare professional or a care coordinator is keeping track on how the patient is doing after treatments end (ex: a structured follow-up care plan).

#### Patient support groups

“A group of people with common experiences and concerns who provide emotional and moral support for one another.”47

#### Home-based care

Care or services that are provided at home, out of the hospital.

#### Telehealth / virtual care

Care or services that are provided over the phone or the Internet (on a computer, etc.).

## APPENDIX 3 Search Strategies

Long Covid – Models, CPGS, SRs

### Final Strategies

2021 May 27

### Ovid Multifile

Database: Ovid MEDLINE: Epub Ahead of Print, In-Process & Other Non-Indexed Citations, Ovid MEDLINE® Daily and Ovid MEDLINE® <1946-Present>, Embase Classic+Embase <1947 to 2021 May 26>

### Search Strategy

1. (long adj (COVID or COVID-19 or COVID19 or coronavirus* or corona virus* or 2019-nCoV or 19nCoV or 2019nCoV or nCoV or n-CoV or “CoV 2” or CoV2 or SARS-CoV-2 or SARS-CoV2 or SARSCoV-2 or SARSCoV2 or SARS2 or SARS-2 or severe acute respiratory syndrome coronavirus 2 or 2019-novel CoV or Sars-coronavirus2 or Sars-coronavirus-2 or SARS-like coronavirus* or novel coronavirus* or novel corona virus* or novel CoV or OC43 or NL63 or 229E or HKU1 or HCoV* or Sars- coronavirus*)).tw,kf. (381)
2. ((longterm or long-term) adj (COVID or COVID-19 or COVID19 or coronavirus* or corona virus* or 2019-nCoV or 19nCoV or 2019nCoV or nCoV or n-CoV or “CoV 2” or CoV2 or SARS-CoV-2 or SARS- CoV2 or SARSCoV-2 or SARSCoV2 or SARS2 or SARS-2 or severe acute respiratory syndrome coronavirus 2 or 2019-novel CoV or Sars-coronavirus2 or Sars-coronavirus-2 or SARS-like coronavirus* or novel coronavirus* or novel corona virus* or novel CoV or OC43 or NL63 or 229E or HKU1 or HCoV* or Sars-coronavirus*)).tw,kf. (41)
3. ((postacute or post-acute) adj (COVID or COVID-19 or COVID19 or coronavirus* or corona virus* or 2019-nCoV or 19nCoV or 2019nCoV or nCoV or n-CoV or “CoV 2” or CoV2 or SARS-CoV-2 or SARS-CoV2 or SARSCoV-2 or SARSCoV2 or SARS2 or SARS-2 or severe acute respiratory syndrome coronavirus 2 or 2019-novel CoV or Sars-coronavirus2 or Sars-coronavirus-2 or SARS-like coronavirus* or novel coronavirus* or novel corona virus* or novel CoV or OC43 or NL63 or 229E or HKU1 or HCoV* or Sars-coronavirus*)).tw,kf. (102)
4. (chronic* adj2 (COVID or COVID-19 or COVID19 or coronavirus* or corona virus* or 2019-nCoV or 19nCoV or 2019nCoV or nCoV or n-CoV or “CoV 2” or CoV2 or SARS-CoV-2 or SARS-CoV2 or SARSCoV-2 or SARSCoV2 or SARS2 or SARS-2 or severe acute respiratory syndrome coronavirus 2 or 2019-novel CoV or Sars-coronavirus2 or Sars-coronavirus-2 or SARS-like coronavirus* or novel coronavirus* or novel corona virus* or novel CoV or OC43 or NL63 or 229E or HKU1 or HCoV* or Sars- coronavirus*)).tw,kf. (151)
5. COVID-19/ and Syndrome/ (91)
6. SARS-CoV-2/ and Syndrome/ (75)
7. or/1-6 [LONG COVID - PT 1] (731)
8. COVID-19/ (83147)
9. SARS-CoV-2/ (74922)
10. Coronavirus/ (13359)
11. Betacoronavirus/ (40862)
12. Coronavirus Infections/ (56621)
13. (COVID-19 or COVID19).tw,kf. (235942)
14. ((coronavirus* or corona virus*) and (hubei or wuhan or beijing or shanghai)).tw,kf. (9410)
15. (wuhan adj5 virus*).tw,kf. (491)
16. (2019-nCoV or 19nCoV or 2019nCoV).tw,kf. (2946)
17. (nCoV or n-CoV or “CoV 2” or CoV2).tw,kf. (85204)
18. (SARS-CoV-2 or SARS-CoV2 or SARSCoV-2 or SARSCoV2 or SARS2 or SARS-2 or severe acute respiratory syndrome coronavirus 2).tw,kf. (86740)
19. (2019-novel CoV or Sars-coronavirus2 or Sars-coronavirus-2 or SARS-like coronavirus* or ((novel or new or nouveau) adj2 (CoV or nCoV or covid or coronavirus* or corona virus or Pandemi*2)) or (coronavirus* and pneumonia)).tw,kf. (34062)
20. (novel coronavirus* or novel corona virus* or novel CoV).tw,kf. (17401)
21. ((coronavirus* or corona virus*) adj2 “2019”).tw,kf. (54774)
22. ((coronavirus* or corona virus*) adj2 “19”).tw,kf. (8784)
23. (coronavirus 2 or corona virus 2).tw,kf. (28182)
24. (OC43 or NL63 or 229E or HKU1 or HCoV* or Sars-coronavirus*).tw,kf. (7411)
25. COVID-19.rx,px,ox. or severe acute respiratory syndrome coronavirus 2.os. (5852)
26. (coronavirus* or corona virus*).ti. (42413)
27. or/8-26 [COVID-19] (294674)
28. (post adj (COVID or COVID-19 or COVID19 or coronavirus* or corona virus* or 2019-nCoV or 19nCoV or 2019nCoV or nCoV or n-CoV or “CoV 2” or CoV2 or SARS-CoV-2 or SARS-CoV2 or SARSCoV-2 or SARSCoV2 or SARS2 or SARS-2 or severe acute respiratory syndrome coronavirus 2 or 2019-novel CoV or Sars-coronavirus2 or Sars-coronavirus-2 or SARS-like coronavirus* or novel coronavirus* or novel corona virus* or novel CoV or OC43 or NL63 or 229E or HKU1 or HCoV* or Sars- coronavirus*) adj3 (comorbid* or “co morbid*” or condition* or convalescen* or disease* or disorder* or illness* or multimorbid* or “multi morbid*” or sickness* or symptom* or syndrome* or sign or signs or prognos* or recuperat* or survivor* or survival* or risk*)).tw,kf. (346)
29. (after adj (COVID or COVID-19 or COVID19 or coronavirus* or corona virus* or 2019-nCoV or 19nCoV or 2019nCoV or nCoV or n-CoV or “CoV 2” or CoV2 or SARS-CoV-2 or SARS-CoV2 or SARSCoV-2 or SARSCoV2 or SARS2 or SARS-2 or severe acute respiratory syndrome coronavirus 2 or 2019-novel CoV or Sars-coronavirus2 or Sars-coronavirus-2 or SARS-like coronavirus* or novel coronavirus* or novel corona virus* or novel CoV or OC43 or NL63 or 229E or HKU1 or HCoV* or Sars- coronavirus*) adj3 (comorbid* or “co morbid*” or condition* or convalescen* or disease* or disorder* or illness* or multimorbid* or “multi morbid*” or sickness* or symptom* or syndrome* or sign or signs or prognos* or recuperat* or survivor* or survival* or risk*)).tw,kf. (330)
30. (following adj (COVID or COVID-19 or COVID19 or coronavirus* or corona virus* or 2019-nCoV or 19nCoV or 2019nCoV or nCoV or n-CoV or “CoV 2” or CoV2 or SARS-CoV-2 or SARS-CoV2 or SARSCoV-2 or SARSCoV2 or SARS2 or SARS-2 or severe acute respiratory syndrome coronavirus 2 or 2019-novel CoV or Sars-coronavirus2 or Sars-coronavirus-2 or SARS-like coronavirus* or novel coronavirus* or novel corona virus* or novel CoV or OC43 or NL63 or 229E or HKU1 or HCoV* or Sars- coronavirus*) adj3 (comorbid* or “co morbid*” or condition* or convalescen* or disease* or disorder* or illness* or multimorbid* or “multi morbid*” or sickness* or symptom* or syndrome* or sign or signs or prognos* or recuperat* or survivor* or survival* or risk*)).tw,kf. (128)
31. ((chronic* or continuous* or continual* or continuing* or delay* or endur* or extend* or fluctuat* or gradual* or lasting* or legacy* or lengthy* or linger* or long* or “medium* term*” or mediumterm* or multisystem* or “multi system*” or ongoing* or permanent* or persist* or prolong* or protract* or relaps* or remission* or remit* or residual* or slow* or subacute* or “sub acute*”) adj3 recover*).tw,kf. (81004)
32. ((after discharg* or following discharg* or postacute* or “post acute*” or postdischarg* or “post discharge” or “post discharging” or posthospital* or post-hospital* or postinfect* or “post infection” or “post infective*” or postviral* or “post viral*” or postvirus* or “post virus*” or postcritical or post-critical or postintensive or post-intensive or post-ICU) adj3 recover*).tw,kf. (1326)
33. ((chronic* or continuous* or continual* or continuing* or delay* or endur* or extend* or fluctuat* or gradual* or lasting* or legacy* or lengthy* or linger* or long* or “medium* term*” or mediumterm* or multisystem* or “multi system*” or ongoing or permanent* or persist* or prolong* or protract* or relaps* or remission* or remit* or residual* or slow* or subacute* or “sub acute*”) adj3 (complication? or consequence? or convalescen* or disabilit* or feature* or illness* or prognos* or sequela* or sign or signs or suffering? or symptom* or recuperat*)).tw,kf. (593443)
34. ((after discharg* or following discharg* or postacute* or “post acute*” or postdischarg* or “post discharge” or “post discharging” or posthospital* or post-hospital* or postinfect* or “post infection” or “post infective*” or postviral* or “post viral*” or postvirus* or “post virus*” or postcritical or post-critical or postintensive or post-intensive or post-ICU) adj3 (complication? or consequence? or convalescen* or disabilit* or feature* or illness* or prognos* or sequela* or sign or signs or suffering? or symptom* or recuperat*)).tw,kf. (4450)
35. (nonrecover* or “non recover*” or “not recover*”).tw,kf. (17092)
36. (“long* haul*” or longhaul* or “long* tail*” or longtail* or longduration* or “long duration*” or longlast* or “long last*” or longstanding* or “long standing*” or “medium* term*” or mediumterm*).tw,kf. (271517)
37. or/28-36 [LONG-TERM ILLNESS, PROTRACTED RECOVERY, ETC.] (942246)
38. 27 and 37 [LONG COVID - PT 2] (6420)
39. Long Term Adverse Effects/ (205340)
40. “Recovery of Function”/ (106473)
41. Convalescence/ (59988)
42. or/39-41 [POST-COVID RECOVERY PERIOD] (320631)
43. 27 and 42 [LONG COVID - PT 3] (2000)
44. 7 or 38 or 43 [LONG COVID] (8693)
45. exp Aftercare/ (1938519)
46. Ambulatory Care/ (85233)
47. Delivery of Health Care/ (244906)
48. Models, Organizational/ (64796)
49. Outpatients/ (138471)
50. exp Patient Care Management/ (1703321)
51. exp Rehabilitation/ (761203)
52. (aftercare or after care or follow-up care).tw,kf. (24545)
53. (rehab or rehabil*).tw,kf. (458901)
54. ((model* or deliver* or framework?) adj5 (care or healthcare or health care or service or services)).tw,kf. (309659)
55. or/45-54 [CARE MODELS] (4750522)
56. 44 and 55 [LONG COVID - CARE MODELS] (1838)
57. exp Clinical Pathways/ (15980)
58. exp Clinical Protocols/ (280688)
59. Consensus/ (93947)
60. exp Consensus Development Conference/ (36866)
61. exp Consensus Development Conferences as Topic/ (27481)
62. exp Guideline/ (35770)
63. Guidelines as Topic/ (458957)
64. Practice Guidelines as Topic/ (475148)
65. Health Planning Guidelines/ (105593)
66. (Guideline or Practice Guideline or Consensus Development Conference or Consensus Development Conference, NIH).pt. (45391)
67. (position statement* or policy statement* or practice parameter* or practice point? or best practice*).tw,kf. (91593)
68. (standards or guidance or guideline or guidelines).ti,kf. (299635)
69. ((practice or treatment* or clinical) adj (guidance or guideline*)).ab. (114984)
70. (CPG or CPGs).ti. (13142)
71. consensus*.ti,kf. (61559)
72. consensus*.ab. /freq=2 (64818)
73. ((care or critical or clinical or practice) adj2 (algorithm* or path or paths or pathway or pathways or protocol*)).tw,kf. (86750)
74. recommendat*.ti,kf. (99351)
75. (overview? adj2 (guidance or guideline?)).tw,kf. (354)
76. exp Clinical Decision-Making/ (63513)
77. ((clinical or medical) adj (decision* or reasoning?)).tw,kf. (109906)
78. or/57-77 [CPG FILTER] (1698463)
79. 44 and 78 [LONG COVID - CPGS] (547)
80. Systematic Review.pt. (155111)
81. exp Systematic Reviews as Topic/ (32120)
82. Meta Analysis.pt. (133028)
83. exp Meta-Analysis as Topic/ (67702)
84. (meta-analy* or metanaly* or metaanaly* or met analy* or integrative research or integrative review* or integrative overview* or research integration or research overview* or collaborative review*).tw,kf. (476868)
85. (systematic review* or systematic overview* or evidence-based review* or evidence-based overview* or (evidence adj3 (review* or overview*)) or meta-review* or meta-overview* or meta- synthes* or rapid review* or “review of reviews” or umbrella review? or technology assessment* or HTA or HTAs).tw,kf. (575494)
86. exp Technology Assessment, Biomedical/ (26395)
87. (cochrane or health technology assessment or evidence report or systematic reviews).jw. (51736)
88. (network adj (MA or MAs)).tw,kf. (32)
89. (NMA or NMAs or MTC or MTCs or MAIC or MAICs).tw,kf. (19824)
90. indirect* compar*.tw,kf. (6193)
91. (indirect treatment* adj1 compar*).tw,kf. (1082)
92. (mixed treatment* adj1 compar*).tw,kf. (1372)
93. (multiple treatment* adj1 compar*).tw,kf. (423)
94. (multi-treatment* adj1 compar*).tw,kf. (9)
95. simultaneous* compar*.tw,kf. (2585)
96. mixed comparison?.tw,kf. (85)
97. or/80-96 [REVIEW FILTER] (966617)
98. 44 and 97 [LONG COVID - REVIEWS] (338)
99. 56 or 79 or 98 [LONG COVID - MODELS OF CARE, CPGS, REVIEWS] (2408)
100. 99 use ppez [MEDLINE RECORDS] (837)
101. (long adj (COVID or COVID-19 or COVID19 or coronavirus* or corona virus* or 2019-nCoV or 19nCoV or 2019nCoV or nCoV or n-CoV or “CoV 2” or CoV2 or SARS-CoV-2 or SARS-CoV2 or SARSCoV-2 or SARSCoV2 or SARS2 or SARS-2 or severe acute respiratory syndrome coronavirus 2 or 2019-novel CoV or Sars-coronavirus2 or Sars-coronavirus-2 or SARS-like coronavirus* or novel coronavirus* or novel corona virus* or novel CoV or OC43 or NL63 or 229E or HKU1 or HCoV* or Sars- coronavirus*)).tw,kw. (375)
102. ((longterm or long-term) adj (COVID or COVID-19 or COVID19 or coronavirus* or corona virus* or 2019-nCoV or 19nCoV or 2019nCoV or nCoV or n-CoV or “CoV 2” or CoV2 or SARS-CoV-2 or SARS-CoV2 or SARSCoV-2 or SARSCoV2 or SARS2 or SARS-2 or severe acute respiratory syndrome coronavirus 2 or 2019-novel CoV or Sars-coronavirus2 or Sars-coronavirus-2 or SARS-like coronavirus* or novel coronavirus* or novel corona virus* or novel CoV or OC43 or NL63 or 229E or HKU1 or HCoV* or Sars-coronavirus*)).tw,kw. (47)
103. ((postacute or post-acute) adj (COVID or COVID-19 or COVID19 or coronavirus* or corona virus* or 2019-nCoV or 19nCoV or 2019nCoV or nCoV or n-CoV or “CoV 2” or CoV2 or SARS-CoV-2 or SARS-CoV2 or SARSCoV-2 or SARSCoV2 or SARS2 or SARS-2 or severe acute respiratory syndrome coronavirus 2 or 2019-novel CoV or Sars-coronavirus2 or Sars-coronavirus-2 or SARS-like coronavirus* or novel coronavirus* or novel corona virus* or novel CoV or OC43 or NL63 or 229E or HKU1 or HCoV* or Sars-coronavirus*)).tw,kw. (104)
104. (chronic* adj2 (COVID or COVID-19 or COVID19 or coronavirus* or corona virus* or 2019-nCoV or 19nCoV or 2019nCoV or nCoV or n-CoV or “CoV 2” or CoV2 or SARS-CoV-2 or SARS-CoV2 or SARSCoV-2 or SARSCoV2 or SARS2 or SARS-2 or severe acute respiratory syndrome coronavirus 2 or 2019-novel CoV or Sars-coronavirus2 or Sars-coronavirus-2 or SARS-like coronavirus* or novel coronavirus* or novel corona virus* or novel CoV or OC43 or NL63 or 229E or HKU1 or HCoV* or Sars- coronavirus*)).tw,kw. (638)
105. or/101-104 [LONG COVID - PT 1] (1121)
106. coronavirus disease 2019/ (193990)
107. severe acute respiratory syndrome coronavirus 2/ (92200)
108. Coronavirinae/ (4840)
109. Betacoronavirus/ (40862)
110. coronavirus infection/ (57497)
111. (COVID-19 or COVID19).tw,kw. (239859)
112. ((coronavirus* or corona virus*) and (hubei or wuhan or beijing or shanghai)).tw,kw. (9550)
113. (wuhan adj5 virus*).tw,kw. (514)
114. (2019-nCoV or 19nCoV or 2019nCoV).tw,kw. (3271)
115. (nCoV or n-CoV or “CoV 2” or CoV2).tw,kw. (84744)
116. (SARS-CoV-2 or SARS-CoV2 or SARSCoV-2 or SARSCoV2 or SARS2 or SARS-2 or severe acute respiratory syndrome coronavirus 2).tw,kw. (92405)
117. (2019-novel CoV or Sars-coronavirus2 or Sars-coronavirus-2 or SARS-like coronavirus* or ((novel or new or nouveau) adj2 (CoV or nCoV or covid or coronavirus* or corona virus or Pandemi*2)) or (coronavirus* and pneumonia)).tw,kw. (34473)
118. (novel coronavirus* or novel corona virus* or novel CoV).tw,kw. (17732)
119. ((coronavirus* or corona virus*) adj2 “2019”).tw,kw. (54567)
120. ((coronavirus* or corona virus*) adj2 “19”).tw,kw. (8528)
121. (coronavirus 2 or corona virus 2).tw,kw. (27983)
122. (OC43 or NL63 or 229E or HKU1 or HCoV* or Sars-coronavirus*).tw,kw. (7585)
123. (coronavirus* or corona virus*).ti. (42413)
124. or/106-123 [COVID-19] (303261)
125. (post adj (COVID or COVID-19 or COVID19 or coronavirus* or corona virus* or 2019-nCoV or 19nCoV or 2019nCoV or nCoV or n-CoV or “CoV 2” or CoV2 or SARS-CoV-2 or SARS-CoV2 or SARSCoV-2 or SARSCoV2 or SARS2 or SARS-2 or severe acute respiratory syndrome coronavirus 2 or 2019-novel CoV or Sars-coronavirus2 or Sars-coronavirus-2 or SARS-like coronavirus* or novel coronavirus* or novel corona virus* or novel CoV or OC43 or NL63 or 229E or HKU1 or HCoV* or Sars- coronavirus*) adj3 (comorbid* or “co morbid*” or condition* or convalescen* or disease* or disorder* or illness* or multimorbid* or “multi morbid*” or sickness* or symptom* or syndrome* or sign or signs or prognos* or recuperat* or survivor* or survival* or risk*)).tw,kw. (347)
126. (after adj (COVID or COVID-19 or COVID19 or coronavirus* or corona virus* or 2019-nCoV or 19nCoV or 2019nCoV or nCoV or n-CoV or “CoV 2” or CoV2 or SARS-CoV-2 or SARS-CoV2 or SARSCoV-2 or SARSCoV2 or SARS2 or SARS-2 or severe acute respiratory syndrome coronavirus 2 or 2019-novel CoV or Sars-coronavirus2 or Sars-coronavirus-2 or SARS-like coronavirus* or novel coronavirus* or novel corona virus* or novel CoV or OC43 or NL63 or 229E or HKU1 or HCoV* or Sars- coronavirus*) adj3 (comorbid* or “co morbid*” or condition* or convalescen* or disease* or disorder* or illness* or multimorbid* or “multi morbid*” or sickness* or symptom* or syndrome* or sign or signs or prognos* or recuperat* or survivor* or survival* or risk*)).tw,kw. (330)
127. (following adj (COVID or COVID-19 or COVID19 or coronavirus* or corona virus* or 2019-nCoV or 19nCoV or 2019nCoV or nCoV or n-CoV or “CoV 2” or CoV2 or SARS-CoV-2 or SARS-CoV2 or SARSCoV-2 or SARSCoV2 or SARS2 or SARS-2 or severe acute respiratory syndrome coronavirus 2 or 2019-novel CoV or Sars-coronavirus2 or Sars-coronavirus-2 or SARS-like coronavirus* or novel coronavirus* or novel corona virus* or novel CoV or OC43 or NL63 or 229E or HKU1 or HCoV* or Sars- coronavirus*) adj3 (comorbid* or “co morbid*” or condition* or convalescen* or disease* or disorder* or illness* or multimorbid* or “multi morbid*” or sickness* or symptom* or syndrome* or sign or signs or prognos* or recuperat* or survivor* or survival* or risk*)).tw,kw. (128)
128. ((chronic* or continuous* or continual* or continuing* or delay* or endur* or extend* or fluctuat* or gradual* or lasting* or legacy* or lengthy* or linger* or long* or “medium* term*” or mediumterm* or multisystem* or “multi system*” or ongoing* or permanent* or persist* or prolong* or protract* or relaps* or remission* or remit* or residual* or slow* or subacute* or “sub acute*”) adj3 recover*).tw,kw. (81353)
129. ((after discharg* or following discharg* or postacute* or “post acute*” or postdischarg* or “post discharge” or “post discharging” or posthospital* or post-hospital* or postinfect* or “post infection” or “post infective*” or postviral* or “post viral*” or postvirus* or “post virus*” or postcritical or post-critical or postintensive or post-intensive or post-ICU) adj3 recover*).tw,kw. (1335)
130. ((chronic* or continuous* or continual* or continuing* or delay* or endur* or extend* or fluctuat* or gradual* or lasting* or legacy* or lengthy* or linger* or long* or “medium* term*” or mediumterm* or multisystem* or “multi system*” or ongoing or permanent* or persist* or prolong* or protract* or relaps* or remission* or remit* or residual* or slow* or subacute* or “sub acute*”) adj3 (complication? or consequence? or convalescen* or disabilit* or feature* or illness* or prognos* or sequela* or sign or signs or suffering? or symptom* or recuperat*)).tw,kw. (597936)
131. ((after discharg* or following discharg* or postacute* or “post acute*” or postdischarg* or “post discharge” or “post discharging” or posthospital* or post-hospital* or postinfect* or “post infection” or “post infective*” or postviral* or “post viral*” or postvirus* or “post virus*” or postcritical or post-critical or postintensive or post-intensive or post-ICU) adj3 (complication? or consequence? or convalescen* or disabilit* or feature* or illness* or prognos* or sequela* or sign or signs or suffering? or symptom* or recuperat*)).tw,kw. (4478)
132. (nonrecover* or “non recover*” or “not recover*”).tw,kw. (17095)
133. (“long* haul*” or longhaul* or “long* tail*” or longtail* or longduration* or “long duration*” or longlast* or “long last*” or longstanding* or “long standing*” or “medium* term*” or mediumterm*).tw,kw. (271747)
134. or/125-133 [LONG-TERM ILLNESS, PROTRACTED RECOVERY, ETC.] (947181)
135. 124 and 134 [LONG COVID - PT 2] (6532)
136. convalescence/ (59988)
137. and 136 [LONG COVID - PT 3] (1236)
138. or 135 or 137 [LONG COVID] (8425)
139. exp aftercare/ (1938519)
140. exp ambulatory care/ (106994)
141. health care delivery/ (281754)
142. nonbiological model/ (48215)
143. outpatient/ (156497)
144. outpatient care/ (84406)
145. patient care/ (324986)
146. patient care planning/ (69401)
147. exp rehabilitation/ (761203)
148. (aftercare or after care or follow-up care).tw,kw. (24870)
149. (rehab or rehabil*).tw,kw. (472388)
150. ((model* or deliver* or framework?) adj5 (care or healthcare or health care or service or services)).tw,kw. (303666)
151. or/139-150 [CARE MODELS] (3802557)
152. 138 and 151 [LONG COVID - CARE MODELS] (1483)
153. exp practice guideline/ (625602)
154. (position statement* or policy statement* or practice parameter* or practice point? or best practice*).tw,kw. (92164)
155. (standards or guidance or guideline or guidelines).ti,kw. (322387)
156. ((practice or treatment* or clinical) adj (guidance or guideline*)).ab. (114984)
157. (CPG or CPGs).ti. (13142)
158. consensus/ (93947)
159. consensus*.ti,kw. (64562)
160. consensus*.ab. /freq=2 (64818)
161. ((care or critical or clinical or practice) adj2 (algorithm* or path or paths or pathway or pathways or protocol*)).tw,kw. (87461)
162. recommendat*.ti,kw. (102225)
163. (overview? adj2 (guidance or guideline?)).tw,kw. (358)
164. clinical decision making/ (63185)
165. ((clinical or medical) adj (decision* or reasoning?)).tw,kw. (110930)
166. or/153-165 [CPG FILTER] (1383950)
167. 138 and 166 [LONG COVID - CPGS] (468)
168. “systematic review”/ (452145)
169. “systematic review (topic)”/ (26590)
170. meta-analysis/ (348764)
171. “meta analysis (topic)”/ (45671)
172. (meta-analy* or metanaly* or metaanaly* or met analy* or integrative research or integrative review* or integrative overview* or research integration or research overview* or collaborative review*).tw,kw. (480250)
173. (systematic review* or systematic overview* or evidence-based review* or evidence-based overview* or (evidence adj3 (review* or overview*)) or meta-review* or meta-overview* or meta- synthes* or rapid review* or “review of reviews” or umbrella review? or technology assessment* or HTA or HTAs).tw,kw. (579797)
174. exp Technology Assessment, Biomedical/ (26395)
175. (cochrane or health technology assessment or evidence report or systematic reviews).jw. (51736)
176. (network adj (MA or MAs)).tw,kw. (32)
177. (NMA or NMAs or MTC or MTCs or MAIC or MAICs).tw,kw. (19881)
178. indirect* compar*.tw,kw. (6293)
179. (indirect treatment* adj1 compar*).tw,kw. (1088)
180. (mixed treatment* adj1 compar*).tw,kw. (1395)
181. (multiple treatment* adj1 compar*).tw,kw. (432)
182. (multi-treatment* adj1 compar*).tw,kw. (9)
183. simultaneous* compar*.tw,kw. (2585)
184. mixed comparison?.tw,kw. (86)
185. or/168-184 [REVIEW FILTER] (1057939)
186. 138 and 185 [LONG COVID - REVIEWS] (378)
187. 152 or 167 or 186 [LONG COVID - MODELS OF CARE, CPGS, REVIEWS] (2084)
188. 187 use emczd [EMBASE RECORDS] (1397)
189. 100 or 188 [BOTH DATABASES] (2234)
190. remove duplicates from 189 (1676) [TOTAL UNIQUE RECORDS]
191. 190 use ppez (825) [MEDLINE UNIQUE RECORDS]
192. 190 use emczd (851) [EMBASE UNIQUE RECORDS]

### Web of Science

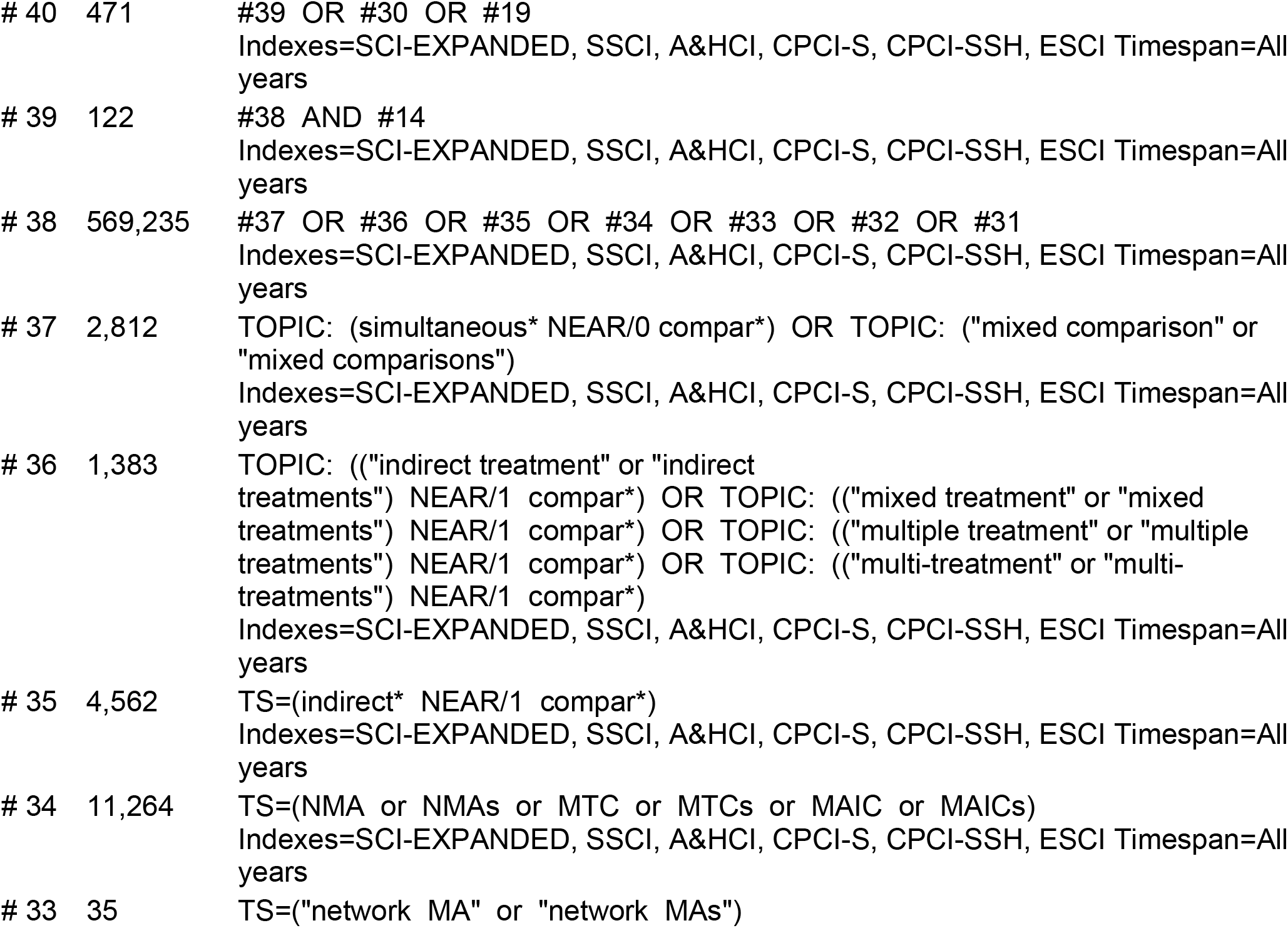
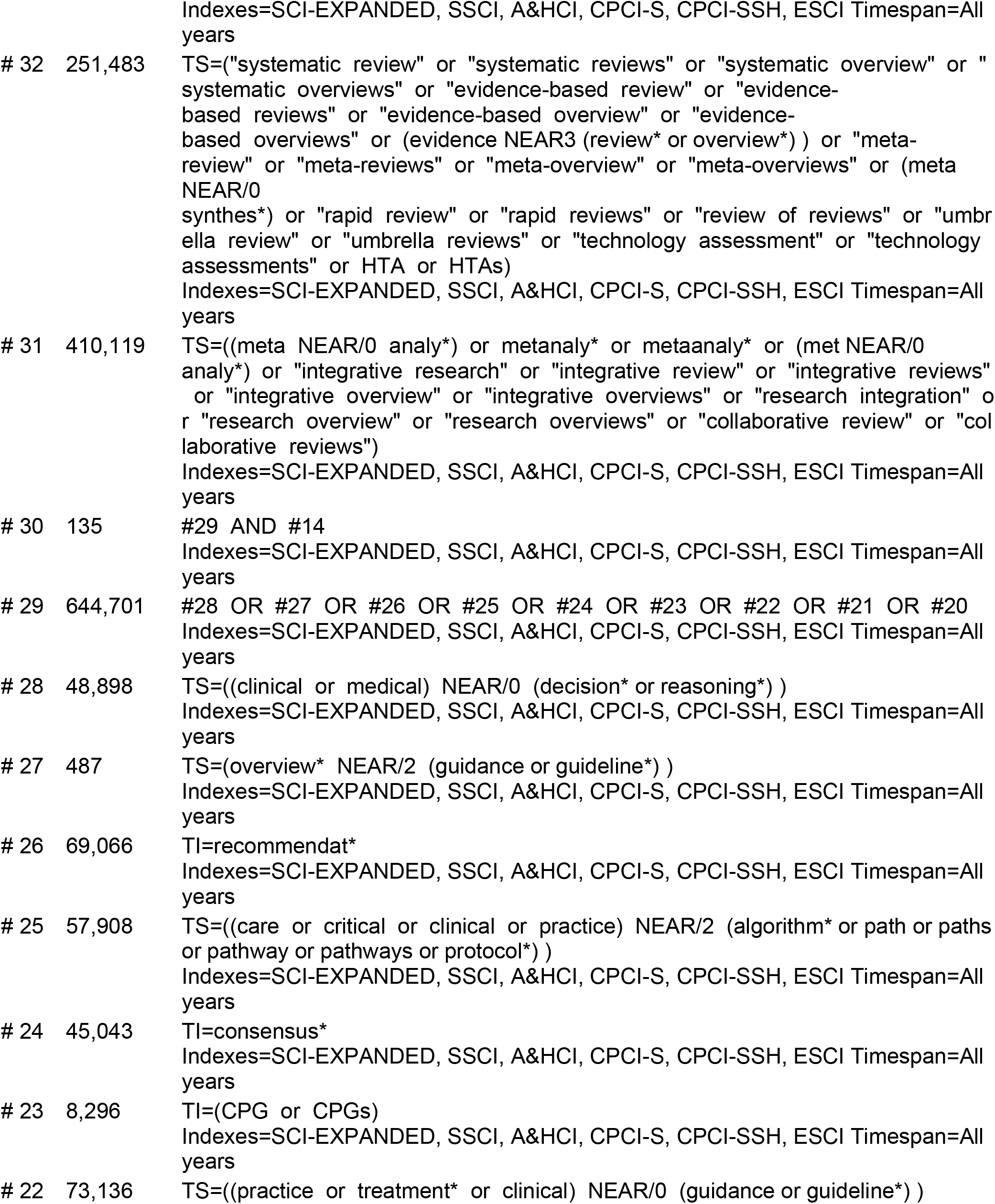
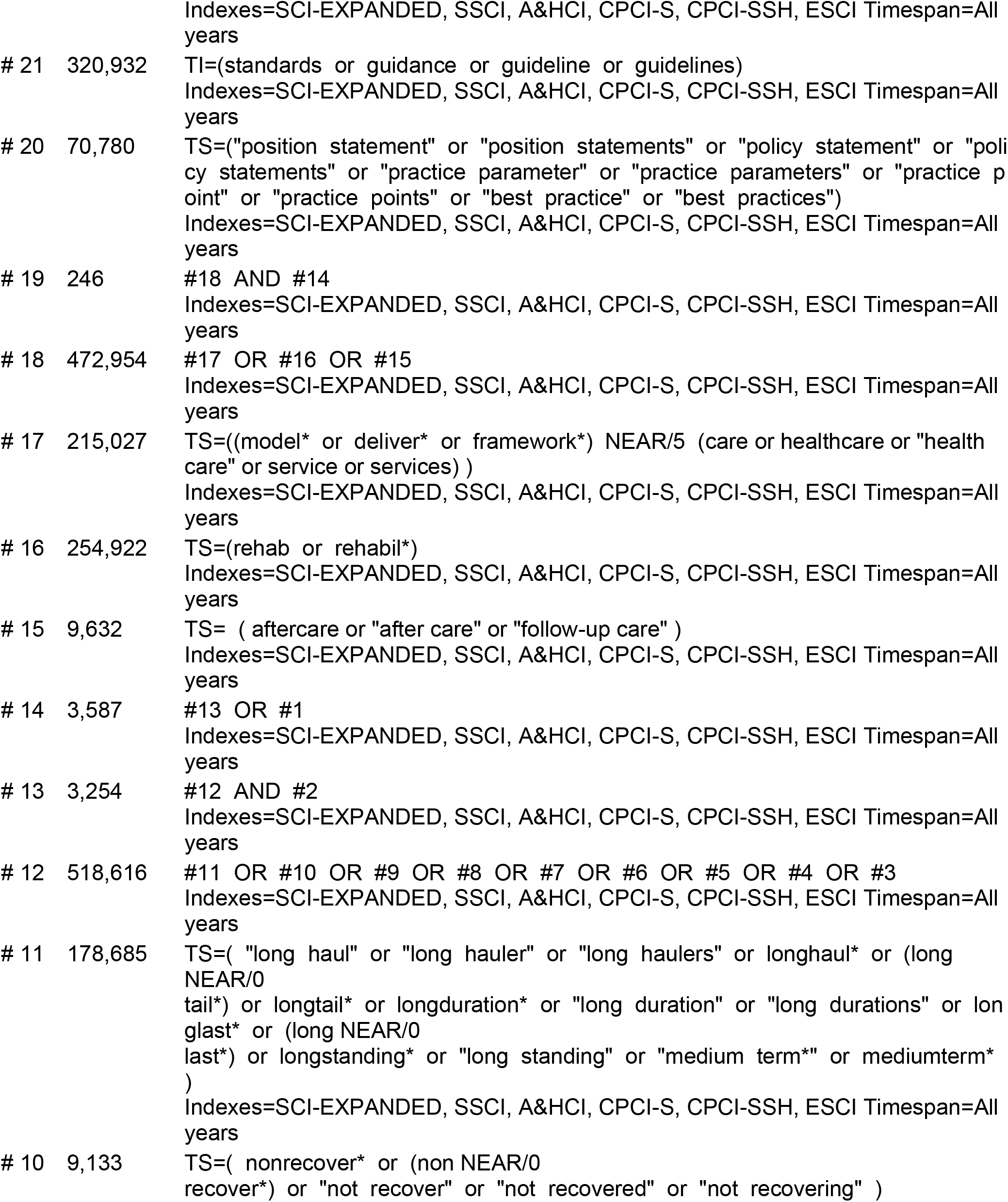
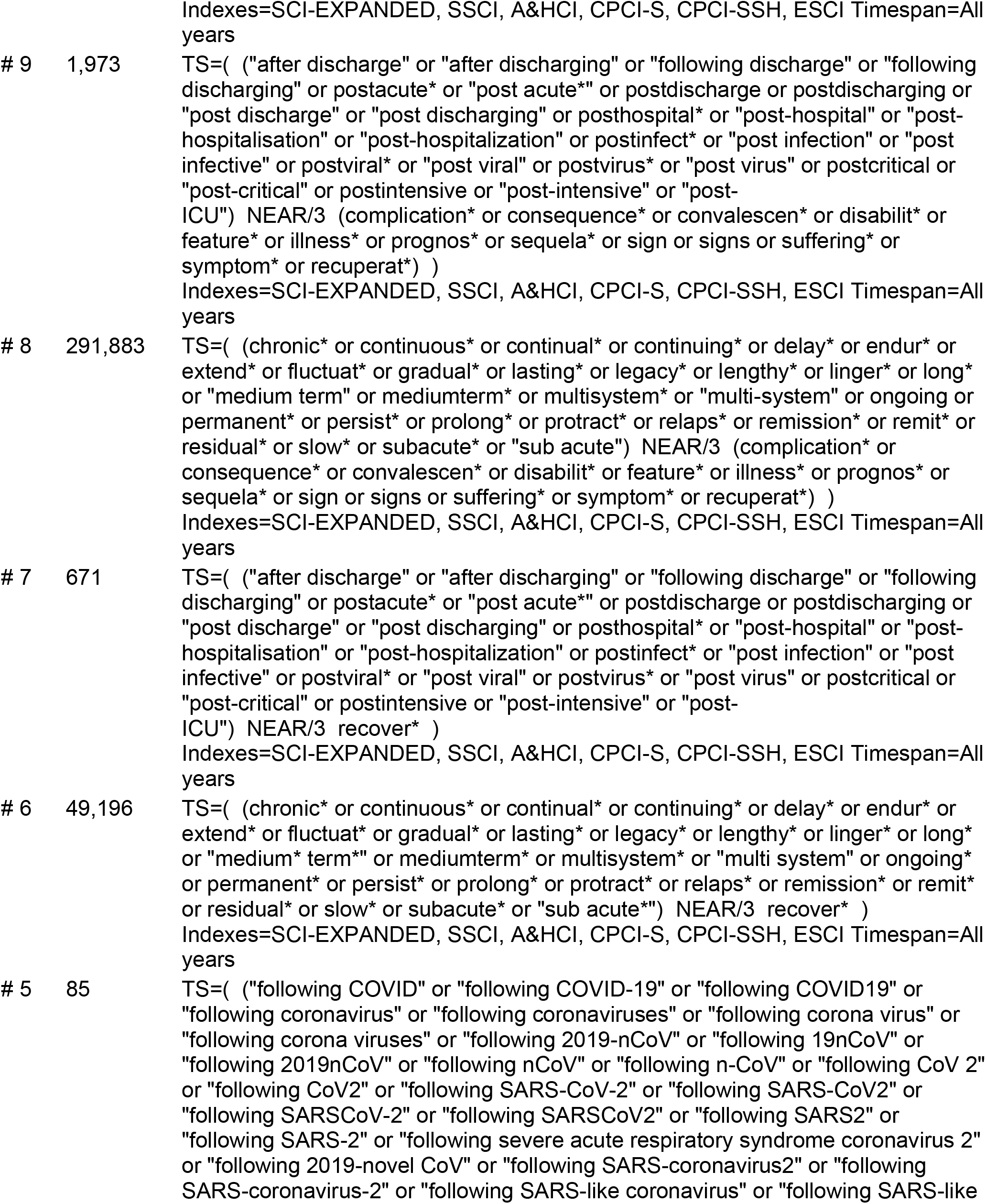
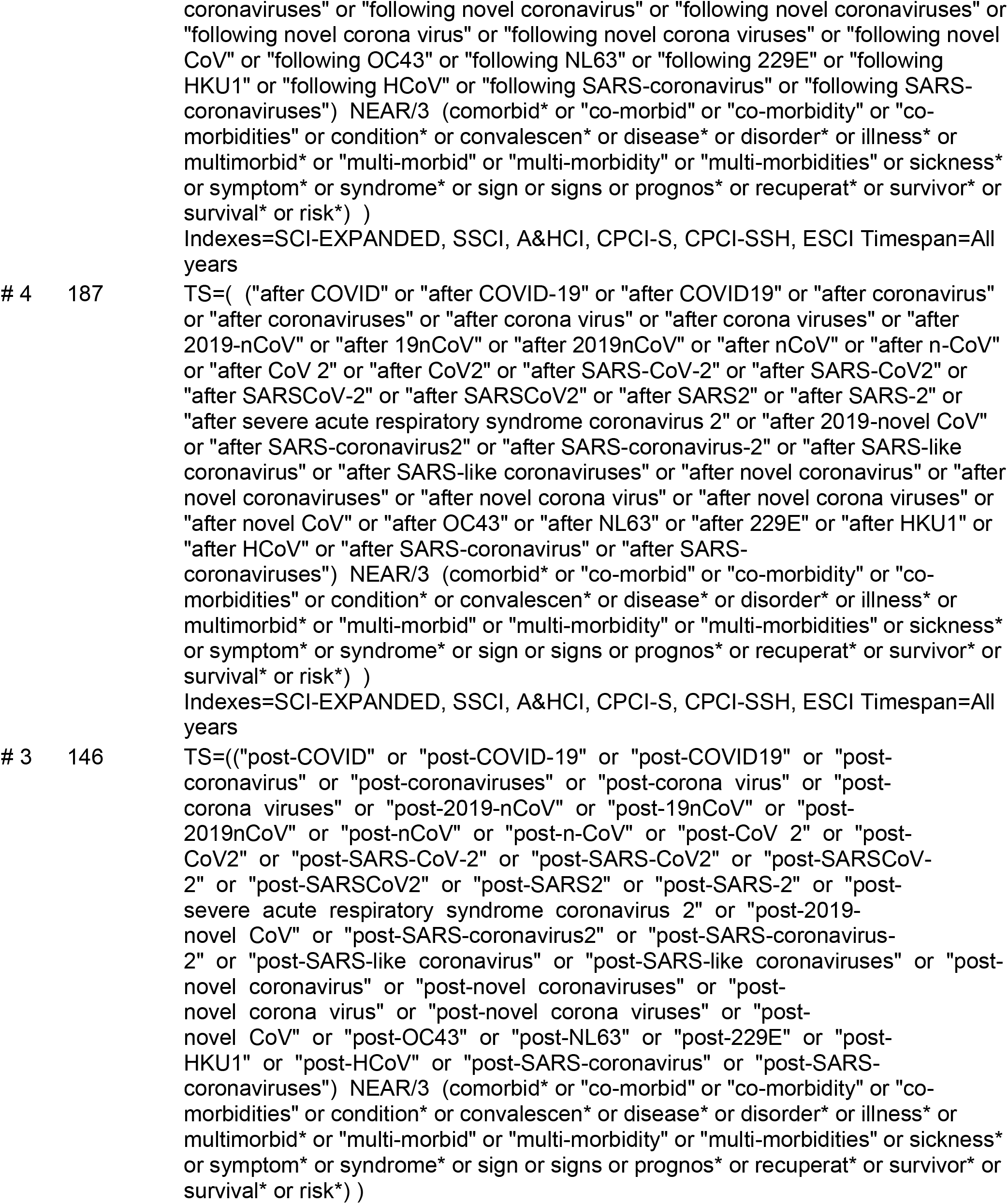
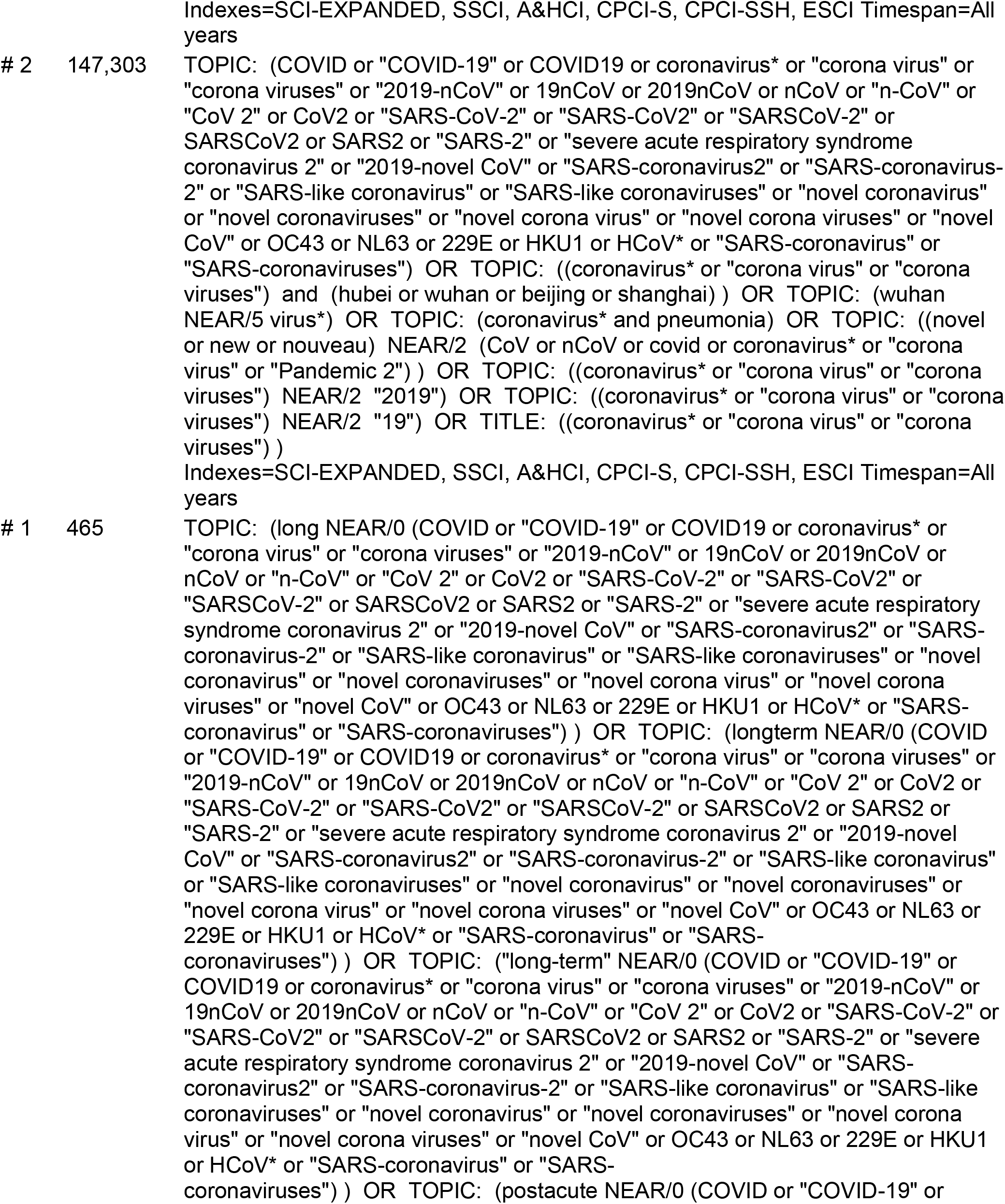
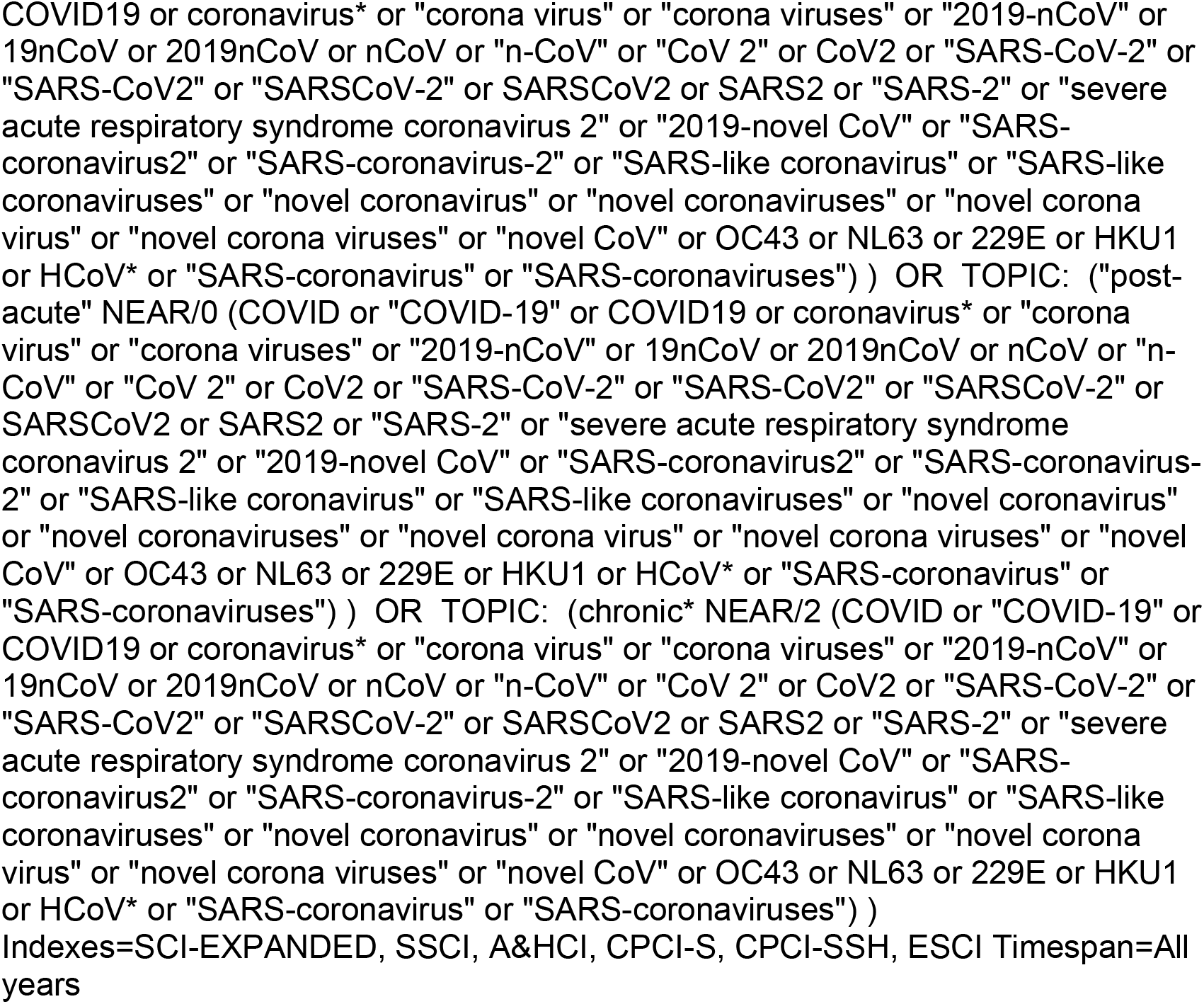

### Cochrane DSR

Search Name:

Date Run: 28/05/2021 02:32:04

Comment:

ID Search Hits

#1 (long NEXT (COVID or “COVID-19” or COVID19 or coronavirus* or “corona virus” or “corona viruses” or “2019-nCoV” or 19nCoV or 2019nCoV or nCoV or “n-CoV” or “CoV 2” or CoV2 or “SARS-CoV-2” or “SARS-CoV2” or “SARSCoV-2” or SARSCoV2 or SARS2 or “SARS-2” or “severe acute respiratory syndrome coronavirus 2” or “2019-novel CoV” or “SARS-coronavirus2” or “SARS-coronavirus-2” or “SARS-like coronavirus” or “SARS-like coronaviruses” or “novel coronavirus” or “novel coronaviruses” or “novel corona virus” or “novel corona viruses” or “novel CoV” or OC43 or NL63 or 229E or HKU1 or HCoV* or “SARS-coronavirus” or “SARS-coronaviruses”)):ti,ab,kw 10

#2 (longterm NEXT (COVID or “COVID-19” or COVID19 or coronavirus* or “corona virus” or “corona viruses” or “2019-nCoV” or 19nCoV or 2019nCoV or nCoV or “n-CoV” or “CoV 2” or CoV2 or “SARS-CoV-2” or “SARS-CoV2” or “SARSCoV-2” or SARSCoV2 or SARS2 or “SARS-2” or “severe acute respiratory syndrome coronavirus 2” or “2019-novel CoV” or “SARS-coronavirus2” or “SARS-coronavirus-2” or “SARS-like coronavirus” or “SARS-like coronaviruses” or “novel coronavirus” or “novel coronaviruses” or “novel corona virus” or “novel corona viruses” or “novel CoV” or OC43 or NL63 or 229E or HKU1 or HCoV* or “SARS-coronavirus” or “SARS-coronaviruses”)):ti,ab,kw 1

#3 (“long-term” NEXT (COVID or “COVID-19” or COVID19 or coronavirus* or “corona virus” or “corona viruses” or “2019-nCoV” or 19nCoV or 2019nCoV or nCoV or “n-CoV” or “CoV 2” or CoV2 or “SARS-CoV-2” or “SARS-CoV2” or “SARSCoV-2” or SARSCoV2 or SARS2 or “SARS-2” or “severe acute respiratory syndrome coronavirus 2” or “2019-novel CoV” or “SARS-coronavirus2” or “SARS-coronavirus-2” or “SARS-like coronavirus” or “SARS-like coronaviruses” or “novel coronavirus” or “novel coronaviruses” or “novel corona virus” or “novel corona viruses” or “novel CoV” or OC43 or NL63 or 229E or HKU1 or HCoV* or “SARS-coronavirus” or “SARS-coronaviruses”)):ti,ab,kw 1

#4 (postacute NEXT (COVID or “COVID-19” or COVID19 or coronavirus* or “corona virus” or “corona viruses” or “2019-nCoV” or 19nCoV or 2019nCoV or nCoV or “n-CoV” or “CoV 2” or CoV2 or “SARS-CoV-2” or “SARS-CoV2” or “SARSCoV-2” or SARSCoV2 or SARS2 or “SARS-2” or “severe acute respiratory syndrome coronavirus 2” or “2019-novel CoV” or “SARS-coronavirus2” or “SARS-coronavirus-2” or “SARS-like coronavirus” or “SARS-like coronaviruses” or “novel coronavirus” or “novel coronaviruses” or “novel corona virus” or “novel corona viruses” or “novel CoV” or OC43 or NL63 or 229E or HKU1 or HCoV* or “SARS-coronavirus” or “SARS-coronaviruses”)):ti,ab,kw 2

#5 (“post-acute” NEXT (COVID or “COVID-19” or COVID19 or coronavirus* or “corona virus” or “corona viruses” or “2019-nCoV” or 19nCoV or 2019nCoV or nCoV or “n-CoV” or “CoV 2” or CoV2 or “SARS-CoV-2” or “SARS-CoV2” or “SARSCoV-2” or SARSCoV2 or SARS2 or “SARS-2” or “severe acute respiratory syndrome coronavirus 2” or “2019-novel CoV” or “SARS-coronavirus2” or “SARS-coronavirus-2” or “SARS-like coronavirus” or “SARS-like coronaviruses” or “novel coronavirus” or “novel coronaviruses” or “novel corona virus” or “novel corona viruses” or “novel CoV” or OC43 or NL63 or 229E or HKU1 or HCoV* or “SARS-coronavirus” or “SARS-coronaviruses”)):ti,ab,kw 2

#6 (chronic* NEAR/2 (COVID or “COVID-19” or COVID19 or coronavirus* or “corona virus” or “corona viruses” or “2019-nCoV” or 19nCoV or 2019nCoV or nCoV or “n-CoV” or “CoV 2” or CoV2 or “SARS-CoV-2” or “SARS-CoV2” or “SARSCoV-2” or SARSCoV2 or SARS2 or “SARS-2” or “severe acute respiratory syndrome coronavirus 2” or “2019-novel CoV” or “SARS-coronavirus2” or “SARS-coronavirus-2” or “SARS-like coronavirus” or “SARS-like coronaviruses” or “novel coronavirus” or “novel coronaviruses” or “novel corona virus” or “novel corona viruses” or “novel CoV” or OC43 or NL63 or 229E or HKU1 or HCoV* or “SARS-coronavirus” or “SARS-coronaviruses”)):ti,ab,kw 6

#7 [or #1-#6] 18

#8 (COVID or “COVID-19” or COVID19 or coronavirus* or “corona virus” or “corona viruses” or “2019-nCoV” or 19nCoV or 2019nCoV or nCoV or “n-CoV” or “CoV 2” or CoV2 or “SARS-CoV-2” or “SARS-CoV2” or “SARSCoV-2” or SARSCoV2 or SARS2 or “SARS-2” or “severe acute respiratory syndrome coronavirus 2” or “2019-novel CoV” or “SARS-coronavirus2” or “SARS-coronavirus-2” or “SARS-like coronavirus” or “SARS-like coronaviruses” or “novel coronavirus” or “novel coronaviruses” or “novel corona virus” or “novel corona viruses” or “novel CoV” or OC43 or NL63 or 229E or HKU1 or HCoV* or “SARS-coronavirus” or “SARS-coronaviruses”):ti,ab,kw 5328

#9 ((coronavirus* or “corona virus” or “corona viruses”) and (hubei or wuhan or beijing or shanghai)):ti,ab,kw 181

#10 (wuhan NEAR/5 virus*):ti,ab,kw 8

#11 (coronavirus* and pneumonia):ti,ab,kw 904

#12 ((novel or new or nouveau) NEAR/2 (CoV or nCoV or covid or coronavirus* or “corona virus” or “Pandemic 2”)):ti,ab,kw 517

#13 ((coronavirus* or “corona virus” or “corona viruses”) NEAR/2 “2019”):ti,ab,kw 1250

#14 ((coronavirus* or “corona virus” or “corona viruses”) NEAR/2 “19”):ti,ab,kw 236

#15 (coronavirus* or “corona virus” or “corona viruses”):ti 628

#16 [or #8-#15] 5328

#17 ((“post-COVID” or “post-COVID-19” or “post-COVID19” or “post-coronavirus” or “post- coronaviruses” or “post-corona virus” or “post-corona viruses” or “post-2019-nCoV” or “post- 19nCoV” or “post-2019nCoV” or “post-nCoV” or “post-n-CoV” or “post-CoV 2” or “post-CoV2” or “post-SARS-CoV-2” or “post-SARS-CoV2” or “post-SARSCoV-2” or “post-SARSCoV2” or “post- SARS2” or “post-SARS-2” or “post-severe acute respiratory syndrome coronavirus 2” or “post- 2019-novel CoV” or “post-SARS-coronavirus2” or “post-SARS-coronavirus-2” or “post-SARS-like coronavirus” or “post-SARS-like coronaviruses” or “post-novel coronavirus” or “post-novel coronaviruses” or “post-novel corona virus” or “post-novel corona viruses” or “post-novel CoV” or “post-OC43” or “post-NL63” or “post-229E” or “post-HKU1” or “post-HCoV” or “post-SARS-coronavirus” or “post-SARS-coronaviruses”) NEAR/3 (comorbid* or “co-morbid” or “co-morbidity” or “co-morbidities” or condition* or convalescen* or disease* or disorder* or illness* or multimorbid* or “multi-morbid” or “multi-morbidity” or “multi-morbidities” or sickness* or symptom* or syndrome* or sign or signs or prognos* or recuperat* or survivor* or survival* or risk*)):ti,ab,kw 10

#18 ((“after COVID” or “after COVID-19” or “after COVID19” or “after coronavirus” or “after coronaviruses” or “after corona virus” or “after corona viruses” or “after 2019-nCoV” or “after 19nCoV” or “after 2019nCoV” or “after nCoV” or “after n-CoV” or “after CoV 2” or “after CoV2” or “after SARS-CoV-2” or “after SARS-CoV2” or “after SARSCoV-2” or “after SARSCoV2” or “after SARS2” or “after SARS-2” or “after severe acute respiratory syndrome coronavirus 2” or “after 2019-novel CoV” or “after SARS-coronavirus2” or “after SARS-coronavirus-2” or “after SARS-like coronavirus” or “after SARS-like coronaviruses” or “after novel coronavirus” or “after novel coronaviruses” or “after novel corona virus” or “after novel corona viruses” or “after novel CoV” or “after OC43” or “after NL63” or “after 229E” or “after HKU1” or “after HCoV” or “after SARS-coronavirus” or “after SARS-coronaviruses”) NEAR/3 (comorbid* or “co-morbid” or “co-morbidity” or “co-morbidities” or condition* or convalescen* or disease* or disorder* or illness* or multimorbid* or “multi-morbid” or “multi-morbidity” or “multi- morbidities” or sickness* or symptom* or syndrome* or sign or signs or prognos* or recuperat* or survivor* or survival* or risk*)):ti,ab,kw 10

#19 ((“following COVID” or “following COVID-19” or “following COVID19” or “following coronavirus” or “following coronaviruses” or “following corona virus” or “following corona viruses” or “following 2019- nCoV” or “following 19nCoV” or “following 2019nCoV” or “following nCoV” or “following n-CoV” or “following CoV 2” or “following CoV2” or “following SARS-CoV-2” or “following SARS-CoV2” or “following SARSCoV-2” or “following SARSCoV2” or “following SARS2” or “following SARS-2” or “following severe acute respiratory syndrome coronavirus 2” or “following 2019-novel CoV” or “following SARS-coronavirus2” or “following SARS-coronavirus-2” or “following SARS-like coronavirus” or “following SARS-like coronaviruses” or “following novel coronavirus” or “following novel coronaviruses” or “following novel corona virus” or “following novel corona viruses” or “following novel CoV” or “following OC43” or “following NL63” or “following 229E” or “following HKU1” or “following HCoV” or “following SARS-coronavirus” or “following SARS-coronaviruses”) NEAR/3 (comorbid* or “co-morbid” or “co-morbidity” or “co-morbidities” or condition* or convalescen* or disease* or disorder* or illness* or multimorbid* or “multi-morbid” or “multi-morbidity” or “multi-morbidities” or sickness* or symptom* or syndrome* or sign or signs or prognos* or recuperat* or survivor* or survival* or risk*)):ti,ab,kw 4

#20 ((chronic* or continuous* or continual* or continuing* or delay* or endur* or extend* or fluctuat* or gradual* or lasting* or legacy* or lengthy* or linger* or long* or “medium* term*” or mediumterm* or multisystem* or “multi system” or ongoing* or permanent* or persist* or prolong* or protract* or relaps* or remission* or remit* or residual* or slow* or subacute* or “sub acute*”) NEAR/3 recover*):ti,ab,kw 4166

#21 ((“after discharge” or “after discharging” or “following discharge” or “following discharging” or postacute* or “post acute*” or postdischarge or postdischarging or “post discharge” or “post discharging” or posthospital* or “post-hospital” or “post-hospitalisation” or “post-hospitalization” or

postinfect* or “post infection” or “post infective” or postviral* or “post viral” or postvirus* or “post virus” or postcritical or “post-critical” or postintensive or “post-intensive” or “post-ICU”) NEAR/3 recover*):ti,ab,kw 137

#22 ((chronic* or continuous* or continual* or continuing* or delay* or endur* or extend* or fluctuat* or gradual* or lasting* or legacy* or lengthy* or linger* or long* or “medium term” or mediumterm* or multisystem* or “multi-system” or ongoing or permanent* or persist* or prolong* or protract* or relaps* or remission* or remit* or residual* or slow* or subacute* or “sub acute”) NEAR/3 (complication* or consequence* or convalescen* or disabilit* or feature* or illness* or prognos* or sequela* or sign or signs or suffering* or symptom* or recuperat*)):ti,ab,kw 30796

#23 ((“after discharge” or “after discharging” or “following discharge” or “following discharging” or postacute* or “post acute*” or postdischarge or postdischarging or “post discharge” or “post discharging” or posthospital* or “post-hospital” or “post-hospitalisation” or “post-hospitalization” or postinfect* or “post infection” or “post infective” or postviral* or “post viral” or postvirus* or “post virus” or postcritical or “post-critical” or postintensive or “post-intensive” or “post-ICU”) NEAR/3 (complication* or consequence* or convalescen* or disabilit* or feature* or illness* or prognos* or sequela* or sign or signs or suffering* or symptom* or recuperat*)):ti,ab,kw 278

#24 (nonrecover* or (non NEAR/0 recover*) or “not recover” or “not recovered” or “not recovering”):ti,ab,kw 385

#25 (“long haul” or “long hauler” or “long haulers” or longhaul* or (long NEAR/0 tail*) or longtail* or longduration* or “long duration” or “long durations” or longlast* or (long NEAR/0 last*) or longstanding* or “long standing” or “medium term*” or mediumterm*):ti,ab,kw 7296

#26 [or #17-#25] 41941

#27 #16 AND #26 179

#28 #7 OR #27 187

### DSR 3

CENTRAL – 183 (*did not download*) Editorial – 1 (*did not download*)

